# PD-RISK Index: Piloting an unsupervised toolkit to identify adults with Parkinson disease

**DOI:** 10.64898/2026.04.30.26352172

**Authors:** Juan Li, Kelsey Grimes, Joseph Saade, David Lewis, Julianna J. Tomlinson, aSCENT-PD Investigators, Andrew Frank, Tim Ramsay, Natalina Salmaso, Douglas Manuel, Michael G. Schlossmacher

## Abstract

Effective screening for Parkinson disease (PD) is important for symptom management and patient recruitment into disease-modifying trials. Here, we examined the performance of the PD-RISK Index toolkit in identifying PD patients at home without supervision.

Between 2022-2024, we recruited 305 participants (ages, 40–85 years): 93 with typical PD; 66 had other neurological conditions; and 146 were healthy controls. Two versions of the toolkit were completed: An original, 69-item-long questionnaire with the 40-scent University-of-Pennsylvania-Smell-Identification-Test (UPSIT); thereafter, a simplified, 11-item-short questionnaire with a newly-developed 8-scent smell test and responses submitted online. The corresponding summary scores (‘PD-RISK Indices’) were calculated for each subject to evaluate diagnostic performances.

In both versions, PD patients had higher questionnaire scores and worse olfaction than healthy controls (*P<0.001*), as expected; scores for subjects with other neurological conditions ranked at intermediate levels. The simplified 11-item questionnaire outperformed the original version in identifying PD. The abbreviated 8-scent smell test and the UPSIT booklet performed equally well in detecting hyposmia. The simplified PD-RISK Index showed the highest diagnostic performance in identifying PD patients amongst all participants; at a value of 23 (range, 0–100, where higher scores indicate higher PD risk), we measured a sensitivity of 0.98, specificity of 0.83, and positive and negative likelihood ratios of 5.88 and 0.02, respectively. The simplified ‘PD-RISK Index’ showed stable performance across sexes and disease duration.

Our study reveals that unsupervised screening for manifest PD can be effectively carried out at home using an 11-item questionnaire, 8-scent smell test and online data entry.

**The plain language title:** The PD-RISK Index study: Facilitating an earlier diagnosis of adults with Parkinson disease by screening at home with a short questionnaire and a simplified smell test

**Plain language summaries:** Effective screening for Parkinson disease (PD) is important for symptom management by healthcare workers and the recruitment of patients into disease modifying trials. We previously published a two-step model for this purpose, which has been validated using several established study cohorts. The study below represents a field-test of this model and the toolkit to implement it (i.e. a self-report questionnaire and a smell test) using a real-world setting.

Between 2022-2024, 305 participants (ages, 40-85 years) were recruited into the study: 93 with typical PD; 66 had other neurological conditions; 146 were healthy. Two versions of the toolkit were completed at-home without supervision by a healthcare worker: (1) an *original*, 69-item-long questionnaire and a 40-scent smell test; (2) a *simplified*, 11-item-short questionnaire with a newly created, 8-scent smell test. Accordingly, two versions of summary scores (‘PD-RISK Index’) were calculated for each subject to examine group classification. Several performance metrics were used to evaluate diagnostic performances and to suggest clinically relevant thresholds.

In both versions, PD patients had significantly higher questionnaire scores (meaning higher risk of having PD) and lower smell test scores (meaning more impaired sense of smell) than healthy controls; scores for subjects with other neurological conditions ranked at intermediate levels. The simplified questionnaire outperformed the original version in diagnostic accuracy. The abbreviated smell test performed as well as the 40-item version in identifying hyposmia. The simplified ‘PD-RISK Index’ showed stable performance across sexes and disease duration.

This study shows that non-invasive and unsupervised screening for manifest PD can be effectively carried out remotely using a simple 11-item questionnaire, an abbreviated 8-scent smell test and an online data entry system, thereby generating a PD-RISK Index for each individual.

## Introduction

Parkinson disease (PD) remains an incurable neurodegenerative disorder. The worldwide pooled prevalence of PD is estimated at 1.51 cases per 1,000 subjects of any age, and 9.34 cases per 1,000 among individuals older than 60 years.^1^ In Canada alone, PD is currently estimated to impact over 110,000 persons directly and to cost $3.3 billion annually; these estimates are expected to grow by 40,000 individuals and an additional $1.1 billion annually by 2034.^2^ Despite the high prevalence and large economic burden, timely access to diagnosis, to early treatment and to care by specialists represents an ongoing challenge; delays negatively impact persons living with PD and their care partners. One potential improvement would be an accurate, readily accessible and inexpensive tool to enable early detection of adults that already have developed symptoms of PD. Such a tool, when implemented within primary care settings, including in remote areas, could help practitioners to establish a working diagnosis of PD, thus enabling them to initiate early symptom management ahead of a formal consultation by a neurologist.^3^ During the last 10 years, several predictive models focusing on PD have been published using carefully curated cohorts launched for the purpose of biomarker research and longitudinal progression studies.^4–11^

The PD-RISK Index approach, especially its original questionnaire, is based on our previously published P_R_EDIGT Score model,^11^ a hypothesis-driven approach to calculate the probability of PD based on epidemiological evidence and known risk categories linked to its pathogenesis. This model had been initially validated using several, established case-control cohorts, including the single-site DeNoPa and multi-centric PPMI studies;^12^ building on it, we designed a 2-step screening approach that paired a standardized questionnaire with an approved smell test (among several biomarkers examined) to test a resource-effective implementation in clinical settings.^12^ This platform has now been re-named by us as ‘PD-RISK Index’ for clarity and to better distinguish it from the ‘PREDICT-PD’ model described by Schrag et al.^6^

Chronic, irreversible hyposmia is a prevalent, non-motor feature associated with PD. However, reduced olfaction can also be associated with cognitive impairment in older adults, including those with no signs of parkinsonism;^13,14^ hence, the causes (and implications) of chronic hyposmia are diverse and intensely researched.^15^ With a focus on PD, we recently derived a simplified smell test that comprises only 8 odorants; it had been generated by using multiple datasets from published cohorts and extensive internal/external validation steps.^16^ As a result, a stand-alone, simplified, scratch-and-sniff smell test was manufactured to facilitate rapid screening for PD-associated hyposmia.

Here, we present the results of a diagnostic classification study, called the PD-RISK Index study, that piloted home-based screening for typical PD in a diverse cohort of subjects contacted after outpatient visits to neurological clinics. Following the study’s completion using the initial tool set^12^ (phase I), we simplified its components^16^ and retested them (phase II). Below, we describe the performance of both versions.

## Materials and methods

The study was conducted and reported in adherence with the STROBE^17^ guideline for observational studies (see **Supplementary Table 8**) and the STARD^18^ guideline for diagnostic accuracy studies (see **Supplementary Table 9**).

### Source of data and participants

The Ottawa, Ontario-based PD-RISK Index study represents a observational, cross-sectional, case-control study that piloted a 2-step diagnostic toolkit by approaching subjects seen at three outpatient clinic sites: The General and Civic campuses of The Ottawa Hospital (TOH; with two sites of a Parkinson’s and Movement Disorders Clinic) and Élisabeth Bruyère Hospital (EBH; Memory Clinic). The two hospitals represent major referral centers in Eastern Ontario for assessment of PD and dementia, respectively. Recruitment and data collection continued over 32 months from February 2022 to September 2024. The study was approved by the Ottawa Health Science Network Research Ethics Board at TOH (REB #: 20210308-01H) and the Research Ethics Board at EBH (REB #: M16-15-050), with participants’ informed consent.

The PD-RISK Index study cohort comprised three diagnostic groups: participants with typical PD [including one subject with dementia with Lewy bodies (DLB)], individuals with other neurological diagnoses (OND), and neurologically healthy controls (HC), all between the ages of 40–85 years at time of enrolment. PD subjects were at Hoehn-Yahr Score I–IV with the majority at II–III (out of V, where a higher score indicates more advanced progression). Based on results of previous validation studies,^12,16^ the originally intended sample size was 50 subjects per group.

Patients were approached at the end of their appointment or via a research database, followed by a written invitation to participate. The HC group comprised patients’ relatives, accompanying persons and unrelated individuals working at or supporting hospital activities. The OND group included cases of atypical parkinsonism, e.g. multiple system atrophy (MSA) and progressive supranuclear palsy (PSP); subjects with mild cognitive impairment (MCI), manifest Alzheimer’s disease (AD), dementias other than AD; and persons with non-neurodegenerative conditions, as listed in detail in **Table 1**. To confirm participants’ diagnoses, electronic medical records were reviewed by three subspecialty-trained neurologists, who were blinded to results of PD-RISK Index assessments. Neurological diagnoses were rendered according to established criteria by the International Movement Disorder Society^19^ and UK Brain Bank^20^ for PD, Consensus Criteria for MSA,^21^ NINDS-PSP Criteria for PSP,^22^ and corresponding criteria for AD ^23^ and MCI.^24^ Diagnostic confirmation was completed for each patient shortly after receiving their responses to the original PD-RISK Index questionnaire.

**Table 1.**
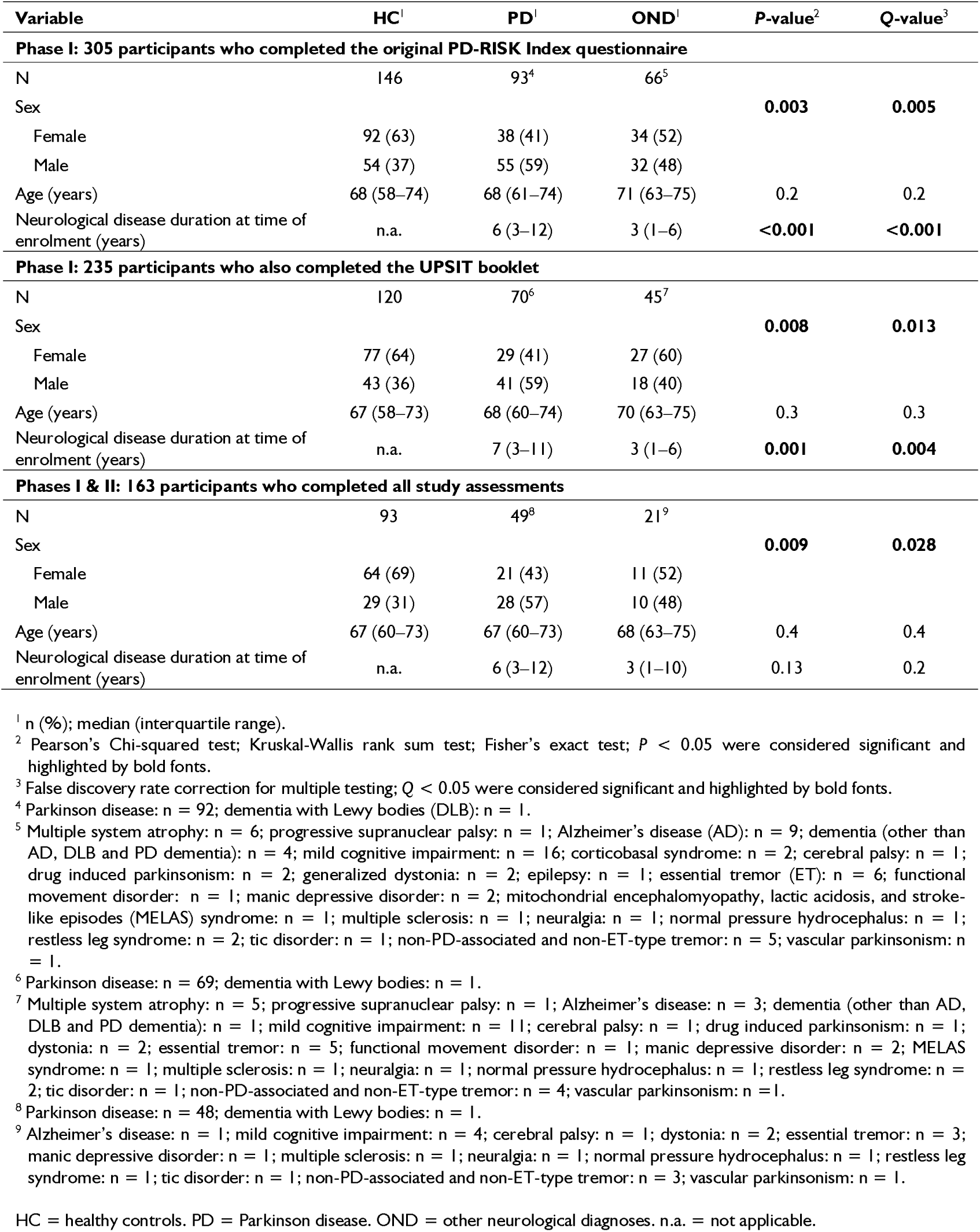
Demographic variables for participants enrolled in the PD-RISK Index study.

| Variable | HC<sup>1</sup> | PD<sup>1</sup> | OND<sup>1</sup> | P-value<sup>2</sup> | Q-value<sup>3</sup> |
| --- | --- | --- | --- | --- | --- |
| **Phase I: 305 participants who completed the original PD-RISK Index questionnaire** |  |  |  |  |  |
| N | 146 | 93<sup>4</sup> | 66<sup>5</sup> |  |  |
| Sex |  |  |  | **0.003** | **0.005** |
| Female | 92 (63) | 38 (41) | 34 (52) |  |  |
| Male | 54 (37) | 55 (59) | 32 (48) |  |  |
| Age (years) | 68 (58–74) | 68 (61–74) | 71 (63–75) | 0.2 | 0.2 |
| Neurological disease duration at time of enrolment (years) | n.a. | 6 (3–12) | 3 (1–6) | **<0.001** | **<0.001** |
| **Phase I: 235 participants who also completed the UPSIT booklet** |  |  |  |  |  |
| N | 120 | 70<sup>6</sup> | 45<sup>7</sup> |  |  |
| Sex |  |  |  | **0.008** | **0.013** |
| Female | 77 (64) | 29 (41) | 27 (60) |  |  |
| Male | 43 (36) | 41 (59) | 18 (40) |  |  |
| Age (years) | 67 (58–73) | 68 (60–74) | 70 (63–75) | 0.3 | 0.3 |
| Neurological disease duration at time of enrolment (years) | n.a. | 7 (3–11) | 3 (1–6) | **0.001** | **0.004** |
| **Phases I & II: 163 participants who completed all study assessments** |  |  |  |  |  |
| N | 93 | 49<sup>8</sup> | 21<sup>9</sup> |  |  |
| Sex |  |  |  | **0.009** | **0.028** |
| Female | 64 (69) | 21 (43) | 11 (52) |  |  |
| Male | 29 (31) | 28 (57) | 10 (48) |  |  |
| Age (years) | 67 (60–73) | 67 (60–73) | 68 (63–75) | 0.4 | 0.4 |
| Neurological disease duration at time of enrolment (years) | n.a. | 6 (3–12) | 3 (1–10) | 0.13 | 0.2 |

### Study assessments and data collection

The study encompassed two phases, in which participants were assessed using the original and a simplified version of the PD-RISK Index toolkit, respectively. **Fig 1** summarizes the workflow.

**Figure 1.**
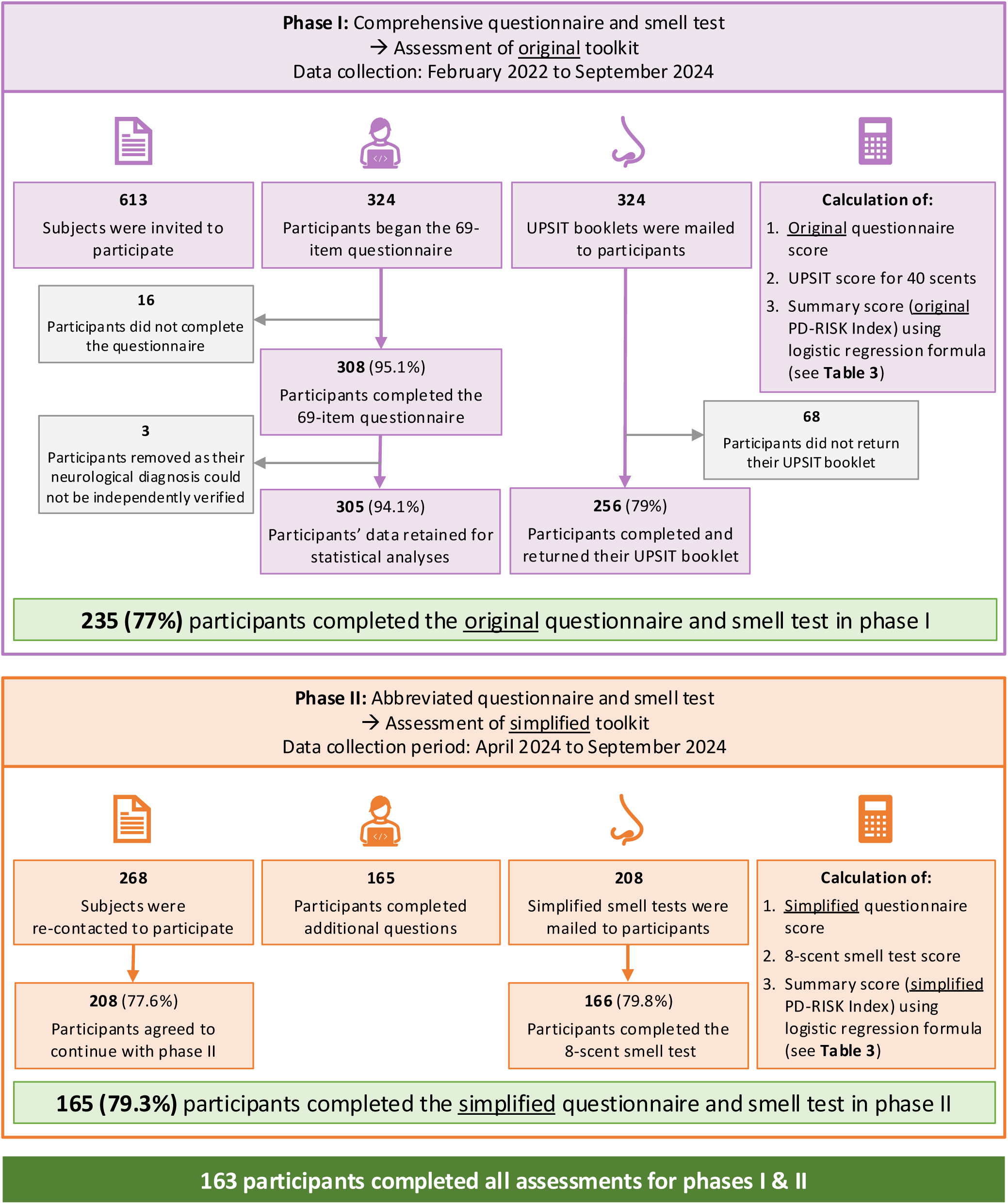
Workflow and data collection summary of the Ottawa-based PD-RISK Index study. UPSIT = University of Pennsylvania Smell Identification Test.

#### Original PD-RISK Index toolkit (phase I)

This version included a comprehensive, self-reported questionnaire hosted on LimeSurvey, which had 69 questions on PD-RISK Index variables^11,12^ (see **Table 2**, **Supplementary Table 1**) and additional ones beyond the original model (not included for score calculation, see **Supplementary Table 2**), i.e. for race/ethnicity, weight/height, physical exercise, medication, and history of select diseases. For the original toolkit, we used the University of Pennsylvania Smell Identification Test (UPSIT)^25^ to assess participants’ olfaction performance: this self-administered kit contains 40 scratch-and-sniff questions for 40 different scents, presented as multiple-choice questions, with 4 options offered for each scent (1 correct option; 3 distractors). UPSIT kits were mailed to participants’ home address with clear instructions; upon completion, marked kits were sent back to the research team for data entry. To eliminate human error in entering responses to 40 UPSIT questions, two research staff members entered data independently, and any discrepancy was resolved by a third person.

**Table 2.**
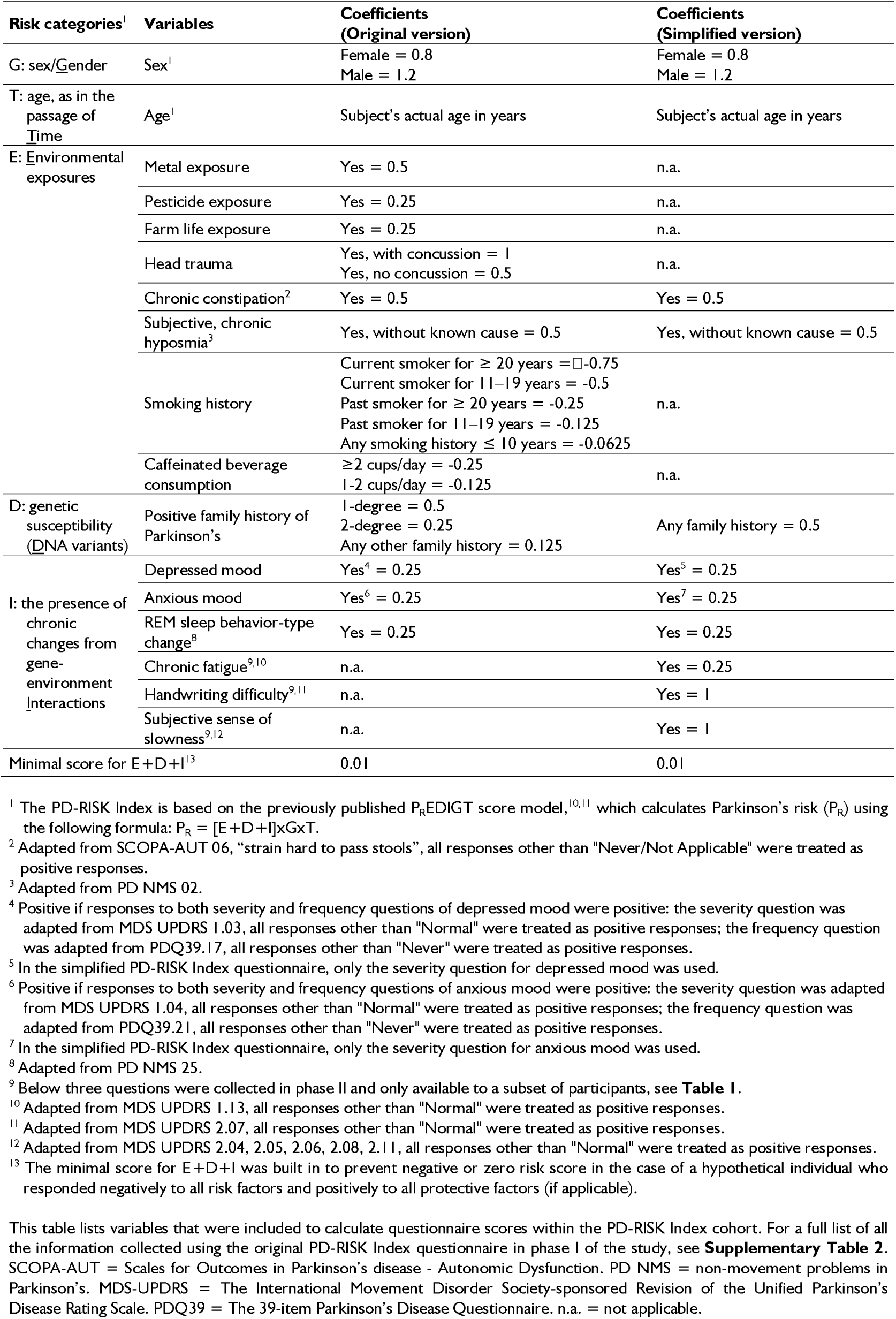
Variables and coefficients to calculate questionnaire scores for the PD-RISK Index.

#### Simplified PD-RISK Index toolkit (phase II)

In this version, the PD-RISK Index questionnaire and associated scoring scheme were updated by adding three carefully selected questions (also self-reported) to capture PD-relevant symptoms (i.e. handwriting difficulty; sense of slowness; fatigue) and by eliminating less informative variables from the original questionnaire (whose odds ratios did not show significance in our previously published validation efforts^12^); the resulting, simplified questionnaire had 11 questions (see **Table 2**). For unsupervised olfaction testing, a simplified, newly developed smell test^16^ (NeuroScent^®^ Card, see **Supplementary Fig 1**) was used: this self-administered card contains 8 scratch-and-sniff odorants, presented as multiple-choice questions, with 5 options offered for each scent: 1 correct option; 3 distractors; and an *“I cannot identify it”* option, which was added to reduce random guessing. Odorants tested in the simplified card version included “dill pickle, mint, rose, coconut, cinnamon, clove, and licorice”. Note, in the current version, one scent was tested twice, with study participants unaware. In phase II, the simplified smell test was mailed to participants’ home address; a separate LimeSurvey questionnaire, completed online by participants at home, was used to collect responses to the newly added questions (i.e. handwriting difficulty; sense of slowness; fatigue) and to the abbreviated smell test. Of note, feasibility and performance of the simplified smell test card was first examined in an unsupervised manner in >150 subjects (separate from those enrolled in the PD-RISK Index study) in the waiting rooms of two hospital-based neurology clinics, where it showed a 99% completion rate for each odorant tested and an ∼10 min completion time (not shown).

### Data preparation and score calculation

The PD-RISK Index questionnaires in phases I and II (hosted on LimeSurvey) could be completed by participants more than once; only the latest, most complete data entry was included for analysis. As illustrated in **Fig 1**, participants needed to complete the original PD-RISK Index questionnaire in phase I to be included for analysis. For UPSIT scoring, invalid responses (blank responses or multiple responses to a single question; <3.5% per question) were assigned 0, indicating an incorrect response. In phase II, all participants except one submitted their responses online and their data were complete without missing entries. One participant only mailed back the completed NeuroScent card but did not respond to the phase II online questionnaire; therefore, only the corresponding simplified smell test score was calculated.

Variables included in the PD-RISK Index questionnaires, their coefficients and the formula (based on the previous P_R_EDIGT Score model) are described in **Table 2**. Coefficients of variables in the original model, and of those remaining in the simplified version, were previously published and remained unchanged.^11,12^ Further, for the three newly added variables (i.e. handwriting difficulty, sense of slowness, fatigue), their coefficients were pre-determined using the same approach as in the original model.^11^ For smell test data (UPSIT and the simplified 8-item test), dichotomous response-based transformation (0 = incorrect; 1 = correct) was used to calculate the sum scores (i.e. number of correctly identified scents). In both phases, the corresponding version of questionnaire scores and smell test scores were integrated using logistic regression to generate a final PD-RISK Index (for details, see **Table 3**). Hence, for each participant as many as 6 scores were generated and compared: (1-1) original questionnaire score, (1-2) UPSIT score, (1-3) original PD-RISK Index; (2-1) simplified questionnaire score, (2-2) simplified smell test score, (2-3) simplified PD-RISK Index.

**Table 3.** Discriminative performances for each test in subjects that completed all assessments in phases I and II.

| Test | Version | Median (interquartile range) |  |  | AUC (95% confidence interval) |  |  |
| --- | --- | --- | --- | --- | --- | --- | --- |
|  |  | HC | PD | OND | PD vs HC | PD vs OND | PD vs HC+OND |
| Scores, Questionnaire<sup>1</sup> | Original<sup>2</sup> | 51<br>(25–82) | 100<br>(71–148) | 82<br>(41–108) | 0.76<br>(0.68, 0.84) | 0.67<br>(0.53, 0.8) | 0.74<br>(0.66, 0.83) |
|  | Simplified<sup>3</sup> | 55<br>(29–82) | 211<br>(170–307) | 93<br>(44–198) | 0.96<br>(0.93, 0.99) | 0.79<br>(0.66, 0.92) | 0.93<br>(0.89, 0.96) |
| Scores, Smell Test | UPSIT<sup>4</sup> | 32<br>(29–35) | 18<br>(13–23) | 27<br>(19–33) | 0.93<br>(0.89, 0.97) | 0.76<br>(0.62, 0.89) | 0.9<br>(0.85, 0.94) |
|  | Simplified<sup>5</sup> | 7<br>(5–8) | 2<br>(1–4) | 6<br>(2–7) | 0.94<br>(0.9, 0.98) | 0.74<br>(0.59, 0.89) | 0.9<br>(0.85, 0.95) |
| **PD-RISK Index** | Original<sup>6</sup> | 7<br>(4–15) | 81<br>(46–95) | 22<br>(7–75) | 0.93<br>(0.89, 0.97) | 0.77<br>(0.65, 0.9) | 0.9<br>(0.86, 0.95) |
|  | Simplified<sup>7</sup> | **2**<br>(1–7) | **97**<br>(70–100) | **14**<br>(2–81) | **0.97**<br>(0.95, 0.99) | **0.8**<br>(0.67, 0.93) | **0.94**<br>(0.91, 0.98) |

<sup>1</sup> Formula to calculate the PD-RISK Index questionnaire score is listed in **Table 2**. For score ranges, the minimum score was based on a hypothetical 40-year-old female who responded negatively to all risk factors and positively to all protective factors (if applicable); the maximum score was based on a hypothetical 100-year-old male who responded positively to all risk factors and negatively to all protective factors (if applicable).
<sup>2</sup> Score range: 0.32–510 (see note 1). Actual score range in the PD-RISK Index study cohort: 0.384–323.7.
<sup>3</sup> Score range: 0.32–540 (see note 1). Actual score range in the PD-RISK Index study cohort: 0.384–398.4.
<sup>4</sup> Score range: 0–40, integer. Actual score range in the PD-RISK Index study cohort: 1–39.
<sup>5</sup> Score range: 0–8, integer. Actual score range in the PD-RISK Index study cohort: 0–8.
<sup>6</sup> Score range: 0–100. Actual score range in the PD-RISK Index study cohort: 0.94–99.89. Predicted probability $\times 100$ based on logistic regression:
<sup>7</sup> Score range: 0–100. Actual score range within the PD-RISK Index study cohort: 0.17–99.99. Predicted probability $\times 100$ based on logistic regression:

The UPSIT booklet is a commonly used smell test in clinical research, while versions of the PD-RISK Index questionnaire and their scoring schemes were developed and pre-determined independently of the current study;^11,12^ therefore, the PD-RISK Index study cohort served as an external validation dataset for these assessments. UPSIT data of a subset of the PD-RISK Index study cohort (n=194/305), along with two other cohorts (n=456), were originally used to develop the simplified smell test;^16^ in phase II, the stand-alone, simplified smell test was completed by 166 study participants (see **Fig 1**); therefore, the PD-RISK Index study cohort served as a mix of temporal and external validation datasets. Further, the current cohort was used to develop the logistic regression models for the two summary scores, where, due to their simple calculation, i.e. linear combination of only two variables (questionnaire score and smell test score; see **Table 3**), overfitting was unlikely. Nonetheless, additional external validation studies using other cohorts are on-going (see Discussion section).

### Data analysis

Demographic and diagnostic characteristics of the study cohort were summarised using n (%) and median (interquartile range). The reported *P*-values represented the significance from corresponding Kruskal-Wallis rank sum test [comparing a continuous or ordinal variable across three groups (i.e. HC, PD, OND)], Pearson’s Chi-squared test (between two categorical variables, with all expected cell counts ≥ 5) or Fisher’s exact test (between two categorical variables, with any expected cell count < 5), with *Q*-values representing false discovery rate correction for multiple testing;^26^ *P*-values and *Q*-values smaller than 0.05 were considered statistically significant.

Univariate analyses reported in **Supplementary Tables 1**,**2**,**4** were used to describe variables collected in the study, both within and beyond the PD-RISK Index model, and to summarize their association with the diagnosis of PD, such as via: number and percentage of positive cases in each group, age- and sex-adjusted odds ratios, and *P*- and *Q*-values for group comparison.

Score distributions in each diagnostic group were illustrated using Cummings estimation plots (e.g. **Fig 2A**),^27^ where a group difference was considered significant when the corresponding confidence interval (shown in the lower panel of each plot) did not cross the zero line. Discrimination performances of these scores to distinguish diagnostic groups (PD vs. HC; PD vs. OND; and PD vs. HC+OND) were compared using area under the receiver operating characteristic curve (AUC) values with bootstrap-estimated 95% confidence intervals (CI)^28^, sensitivity, specificity, positive / negative likelihood ratios, and positive / negative predictive values (as per Shreffler & Huecker^29^). Optimal thresholds reported in **Table 4**, **Supplementary Tables 6**, **7** were determined using the maximum Youden indices.^30^

**Figure 2.**
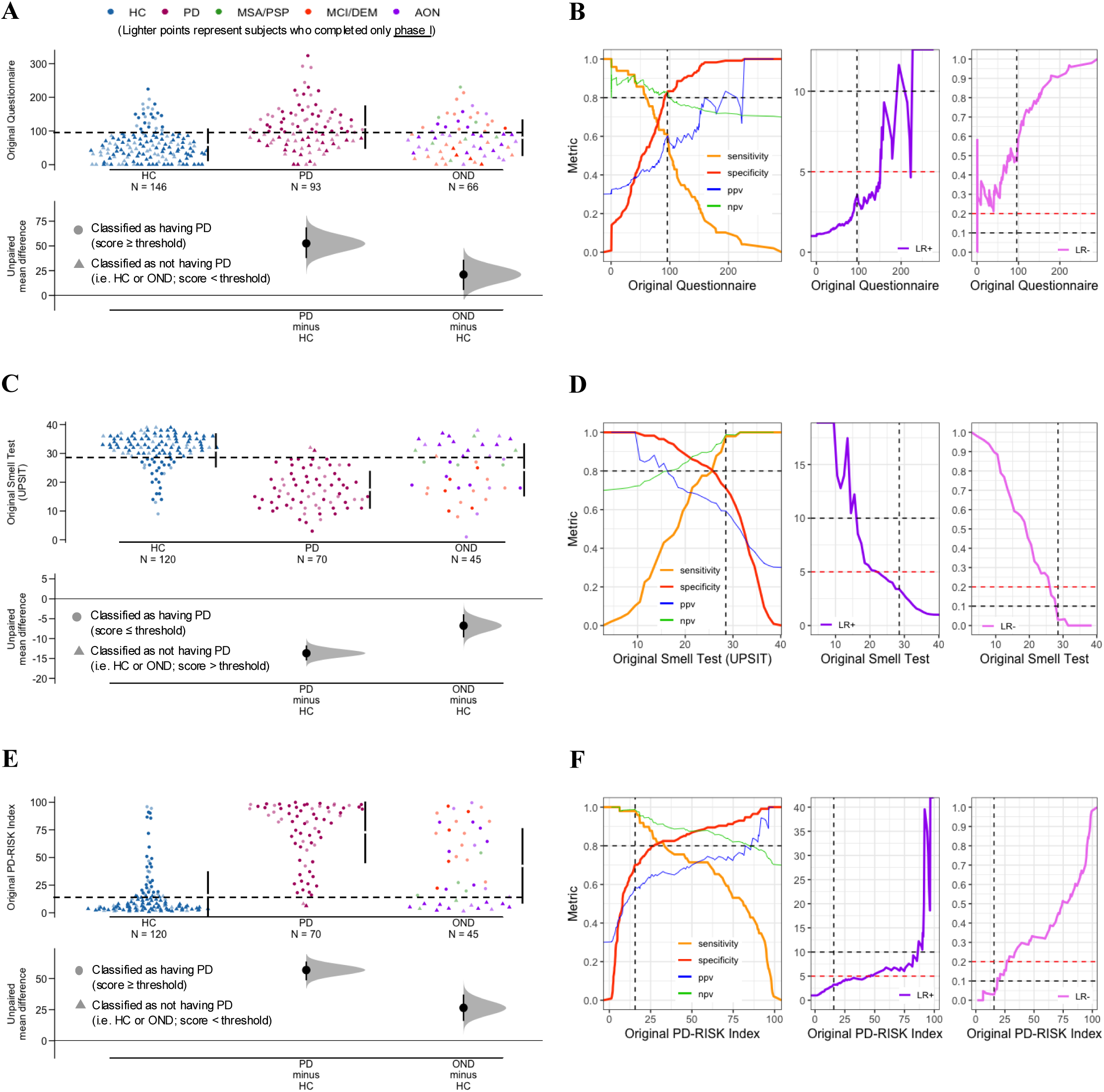
Performances of the original PD-RISK toolkit for three diagnostic groups in the Ottawa-based study. The top (**A**, **B**), middle (**C**, **D**) and bottom (**E**, **F**) rows represent questionnaire score, smell test score and PD-RISK Index, respectively. Panels (**A**, **C**, **E**) illustrate score distributions for three diagnostic groups using Cummings estimation plots. Each symbol represents the score of one participant, and colors identify different groups and diagnoses, as shown in legends in panel (**A**) (for details, see **Table 1**). Horizontal, dashed lines represent the corresponding optimal thresholds for PD vs. HC+OND (see **Table 4**); participants were represented by circular data points when classified as having PD; otherwise, they were represented as data points with triangles. Data points in lighter colors represent participants who completed tests only in phase I. Vertical lines in the upper panel of each graph represent the conventional mean ± standard deviation. Lower panels in each graph show the mean group difference (i.e. effect size) and its 95% confidence interval estimated by bias-corrected and accelerated bootstrapping, using healthy controls as the reference group. Panels (**B**, **D**, **F**) display various performance metrics for the comparison of PD vs. HC+OND (see graph legend). The vertical dashed line in each figure represents the corresponding optimal threshold as shown in **Table 4**. Horizontal dashed lines represent various reference values: sensitivity, specificity, PPV, and NPV values at 0.8; LR+ values at 5 (red) and 10 (black); and LR- values at 0.2 (red) and 0.1 (black). HC = healthy controls. PD = Parkinson disease. OND = other neurological diseases. MSA = multiple system atrophy. PSP = progressive supranuclear palsy. MCI = mild cognitive impairment. DEM = dementia. AON = all other neurological diseases. UPSIT = University of Pennsylvania Smell Identification Test. PPV = positive predictive value. NPV = negative predictive value. LR+ = positive likelihood ratio. LR- = negative likelihood ratio.

**Table 4.** Performance metrics including thresholds for diagnostic classification in the PD-RISK Index study.

| Test | Version | Threshold<sup>1</sup> | Sensitivity | Specificity | LR+ | LR- | PPV | NPV |
| --- | --- | --- | --- | --- | --- | --- | --- | --- |
| **PD vs HC+OND** |  |  |  |  |  |  |  |  |
| Scores, Questionnaire | Original | $\geq 95.83$ | 0.61 | 0.82 | 3.49 | 0.47 | 0.6 | 0.83 |
| | Simplified | $\geq 93.3$ | 0.98 | 0.75 | 3.85 | 0.03 | 0.62 | 0.99 |
| Scores, Smell Test | UPSIT | $\leq 28.5$ | 0.98 | 0.71 | 3.38 | 0.03 | 0.59 | 0.99 |
| | Simplified | $\leq 4.5$ | 0.9 | 0.78 | 4.09 | 0.13 | 0.64 | 0.95 |
| **PD-RISK Index** | Original | $\geq 15.67$ | 0.98 | 0.7 | 3.28 | 0.03 | 0.59 | 0.99 |
|  | **Simplified** | **$\geq 22.72$** | **0.98** | **0.83** | **5.88** | **0.02** | **0.72** | **0.99** |
| **PD vs HC** |  |  |  |  |  |  |  |  |
| Scores, Questionnaire | Original | $\geq 95.83$ | 0.61 | 0.85 | 4.07 | 0.46 | 0.6 | 0.7 |
| | Simplified | $\geq 90.9$ | 1 | 0.78 | 4.65 | 0 | 0.6 | 0.89 |
| Scores, Smell Test | UPSIT | $\leq 28.5$ | 0.98 | 0.76 | 4.14 | 0.03 | 0.59 | 0.87 |
| | Simplified | $\leq 4.5$ | 0.9 | 0.84 | 5.57 | 0.12 | 0.64 | 0.83 |
| **PD-RISK Index** | Original | $\geq 15.67$ | 0.98 | 0.76 | 4.14 | 0.03 | 0.59 | 0.88 |
|  | **Simplified** | **$\geq 22.72$** | **0.98** | **0.89** | **9.11** | **0.02** | **0.72** | **0.86** |
| **PD vs OND** |  |  |  |  |  |  |  |  |
| Scores, Questionnaire | Original | $\geq 90.1$ | 0.63 | 0.71 | 2.21 | 0.51 | 0.56 | 0.14 |
| | Simplified | $\geq 93.3$ | 0.98 | 0.52 | 2.06 | 0.04 | 0.62 | 0.13 |
| Scores, Smell Test | UPSIT | $\leq 28.5$ | 0.98 | 0.48 | 1.87 | 0.04 | 0.59 | 0.12 |
| | Simplified | $\leq 5.5$ | 0.98 | 0.52 | 2.06 | 0.04 | 0.57 | 0.14 |
| **PD-RISK Index** | Original | $\geq 25.62$ | 0.88 | 0.57 | 2.05 | 0.21 | 0.65 | 0.12 |
|  | **Simplified** | **$\geq 19.56$** | **0.98** | **0.57** | **2.29** | **0.04** | **0.71** | **0.13** |

<sup>1</sup> Optimal thresholds are based on the maximum Youden Index value. Signs ( ≥ or ≤ ) indicate classification of 'having PD'.
HC = healthy controls. PD = Parkinson disease. OND = other neurological diagnoses. LR+ = positive likelihood ratio. LR- = negative likelihood ratio. PPV = positive predictive value. NPV = negative predictive value. For explanation and interpretation of these metrics, see Shreffler & Huecker.<sup>28</sup>

Since the PD-RISK Index cohort was insufficiently sex-matched and patients had various lengths of disease at the time of assessment, sub-analyses were performed: effect sizes of PD-RISK Index variables when separating females and males were summarized; performance metrics for all scores were also compared within each sex to exclude bias toward one sex; further, sex-specific thresholds were established in group classification. Comparisons were also made to test effects of disease duration. Further, a relationship between subjective and objective hyposmia was explored by the distribution of smell test scores based on participants’ responses to the question about perceived hyposmia, with or without a known cause.

Analyses were performed using ‘R’ (version 4.5.0) with main packages: ‘gtsummary’,^26^ ‘dabestr’,^27^ ‘pROC’,^28^ and ‘tidyverse^’^.^31^

## Results

### Recruitment into the Ottawa-based PD-RISK Index study cohort

The workflow of research activities and the corresponding number of completions are summarized in **Fig 1**. Among 613 subjects contacted, 324 expressed interest and began answering the questionnaire in phase I; 308 (95.1%) subjects completed all 69 questions. After removing three patients, whose neurological diagnoses could not be independently confirmed, 305 (94.1%) individuals remained for data analysis. Among 324 UPSIT booklets mailed to participants’ home addresses, 256 (79%) were completed and mailed back for data entry. A total of 235 (77%) participants completed all phase I assessments. In phase II, 268 participants, who had been enrolled in phase I, were recontacted. Among these, 208 (77.6%) subjects agreed in writing to continue participating. There, 165 (79.3%) participants completed all phase II assessments and submitted their responses online. By the time of data analysis (January 2025), 163 participants had completed all assessments in both phases of the PD-RISK Index study; none of these were invasive. There was no technical difficulty, discomfort or adverse event reported by study participants.

### Demographic and diagnostic characteristics of the study cohort

The cohort’s demographic and diagnostic characteristics are summarized in **Table 1** and **Supplementary Table 2**. The initial cohort of 305 participants comprised 93 patients with PD (including one patient with DLB), 66 subjects with other neurological conditions, and 146 neurologically healthy controls. The PD and HC groups were age matched with a median age of 68 years, while the OND group was slightly older with a median age of 71 years (*P=0.2*). Sex was not matched, as the percentages of female participants were 63%, 41%, and 52% within HC, PD and OND groups, respectively (*P=0.003*). Patients with PD and OND ranged across disease stages, with median disease duration at 6 and 3 years, respectively (*P<0.001*). Details for subjects that completed all phase I and phase II assessments are listed in **Table 1**; compared to the initial cohort in phase I, no significant changes in overall demographic characteristics were observed in those 163 participants who also completed phase II; however, none of the MSA and PSP subjects completed phase II.

Two participants (both PD patients) identified their ethnicity as being Hispanic or LatinX. Regarding self-identification of race and ethnicity, most of the cohort (over 85% in each group) identified themselves as White (for additional details, see **Supplementary Table 2**).

### Performance of variables collected by two questionnaires

**Table 2** lists variables included in the two versions of the questionnaire and their corresponding coefficients; variables’ effect sizes are listed in **Supplementary Table 1**. When comparing PD vs. HC, effects observed within the PD-RISK Index study cohort were in line with published meta-analyses results for most variables,^3,11^ with the exceptions of pesticide exposure and head trauma (although without statistical significance). Among variables in the original questionnaire, residing on a farm in the past, a history of constipation, subjective olfaction reduction, the presence of depressed mood, and a history of REM-sleep behavior disorder (RBD) were significantly associated with elevated risk of having PD as shown by their corresponding age- and sex-adjusted odds ratios. Further, the three variables added to the simplified questionnaire (i.e. fatigue; handwriting difficulty; sense of slowness) also showed significant association with PD, as expected from their routine inclusion in standardized clinical assessment sheets [e.g. parts I and II of the Unified Parkinson’s Disease Rating Scale (MDS UPDRS)].

Similar associations were also observed when comparing PD vs. OND groups: for most variables, percentages of positive cases within the OND group were at an intermediate level between that of the HC and PD groups. Nonetheless, a history of constipation, subjective olfaction reduction, a positive family history of PD, the presence of RBD, fatigue, handwriting difficulty, and a sense of slowness were more commonly observed among patients with PD than those with other neurological diagnoses. A detailed summary of all variables collected in the study, including those that were not included in the final toolkit, are listed in **Supplementary Table 2**.

### Performance comparison of the original vs. simplified toolkit

The score distribution of original and simplified versions of the toolkit are shown in **Figs 2** and **3**, respectively. The results showed a high degree of consistency between the two versions in that, when compared with the HC group, the PD group had significantly higher questionnaire scores (indicating a higher risk of PD; **Figs 2A**, **3A**) and significantly lower smell tests scores (indicating reduced olfaction; **Figs 2C**, **3C**). The corresponding PD-RISK Index scores, which combined the two assessments, further amplified the significant difference between PD and HC individuals (see **Figs 2E**, **3E**).

**Figure 3.**
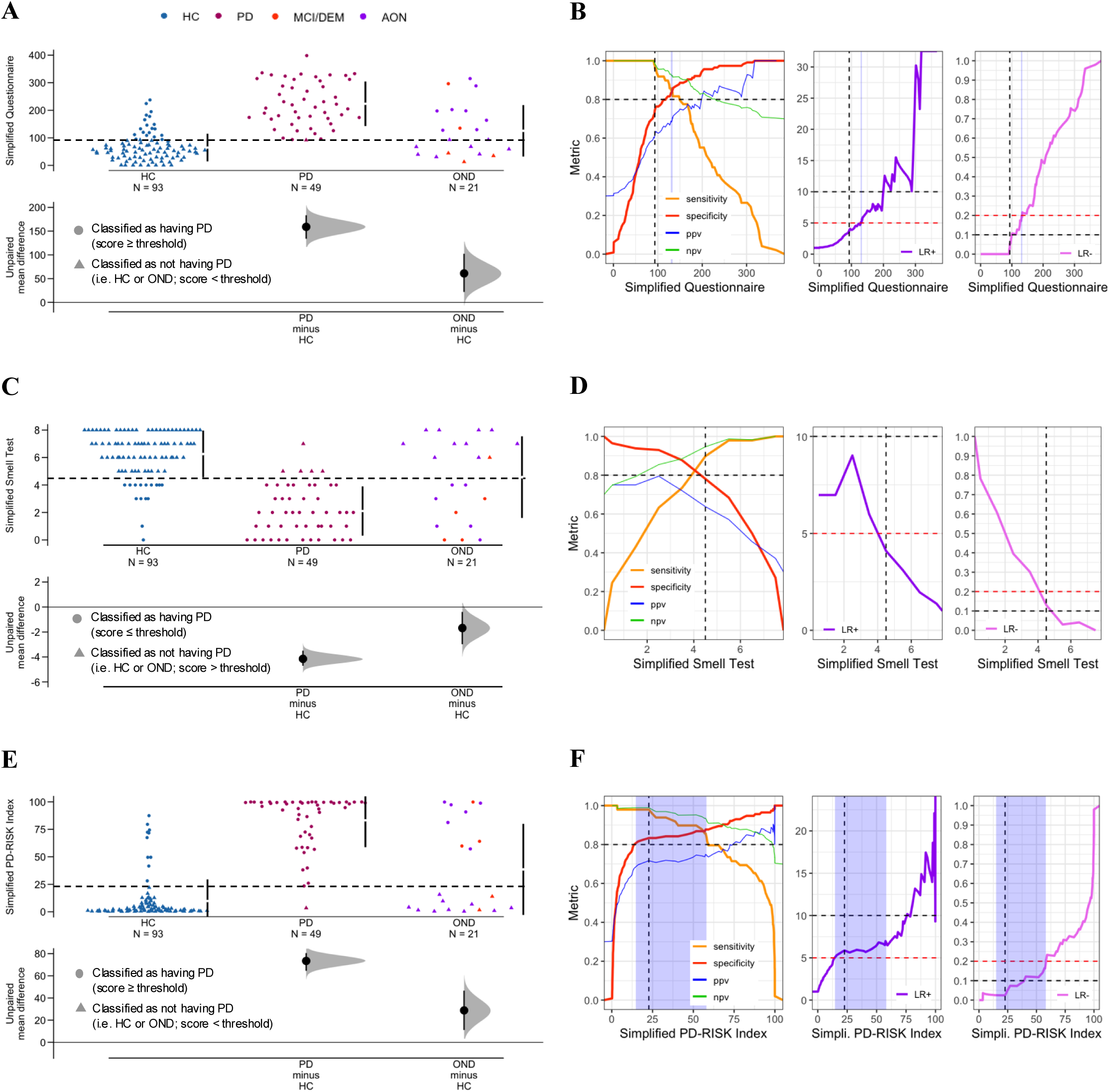
Performances of the simplified PD-RISK toolkit for three diagnostic groups in the Ottawa-based study. The top (**A**, **B**), middle (**C**, **D**) and bottom (**E**, **F**) rows represent questionnaire score, smell test score and PD-RISK Index, respectively. Panels (**A**, **C**, **E**) illustrate score distributions for three diagnostic groups using Cummings estimation plots. Each symbol represents the score of one participant, and colors identify different groups and diagnoses, as shown in legends in panel (**A**) (for details, see **Table 1**). Horizontal, dashed lines represent the corresponding optimal thresholds for PD vs. HC+OND (see **Table 4**); participants were represented by circular data points when classified as having PD; otherwise, they were identified by data points with triangles. Vertical lines in the upper panel of each graph represent the conventional mean ± standard deviation. Lower panels in each graph show the mean group difference (i.e. effect size) and its 95% confidence interval estimated by bias-corrected and accelerated bootstrapping, using healthy controls as the reference group. Panels (**B**, **D**, **F**) display various performance metrics for the comparison of PD vs. HC+OND (see graph legend). The vertical dashed line in each figure represents the corresponding optimal threshold as shown in **Table 4**. Horizontal dashed lines represent various reference values: sensitivity, specificity, PPV, and NPV values at 0.8; LR+ values at 5 (red) and 10 (black); and LR- values at 0.2 (red) and 0.1 (black). The shaded (light blue) areas (in panels **B**, **F**) represent thresholds that satisfy both LR+ > 5 and LR- < 0.2. HC = healthy controls. PD = Parkinson disease. OND = other neurological diseases. MCI = mild cognitive impairment. DEM = dementia. AON = all other neurological diseases. PPV = positive predictive value. NPV = negative predictive value. LR+ = positive likelihood ratio. LR- = negative likelihood ratio.

Scores of the OND group consistently ranked at intermediate levels. There, the MCI/dementia subgroup in our cohort showed a similar degree of hyposmia as the PD group, which is consistent with the literature (see orange symbols in **Figs 2C**, **3C**). Of note, the MSA/PSP subgroup showed relatively better olfaction performance on average when compared with the PD group, as previously shown,^16^ although their sample size was rather small (see green symbols in **Fig 2C**).

When comparing versions of toolkit components, the separation between PD vs. HC groups was further broadened by the simplified questionnaire (**Figs 2A**, **3A**) and, therefore, in the corresponding PD-RISK Index scores (**Figs 2E**, **3E**); this effect was likely due to the inclusion of the three new questions in phase II. Note, as shown in **Fig 2**, we observed no significant difference between participants that only completed phase I (lighter points) vs. those that completed both phases of the study (darker points), which eliminated the possibility of a selection bias for participants in phase II. These findings were supported by group median scores and AUC values for diagnostic classification, as shown in **Table 3**. Scores of the simplified questionnaire and PD-RISK Index outperformed their original counterparts for PD vs. HC, PD vs. OND and PD vs. HC+OND comparisons. Further, with only 8 odorants examined, the simplified smell test card showed a similar discriminative performance to the 40-scent UPSIT kit in identifying subjects with olfaction deficits (**Figs 2C**, **3C**, **Table 3**). Among the 6 scores compared, the simplified PD-RISK Index showed the best performance in accurate group classification, with AUC values of: 0.97 (95%CI 0.95, 0.99) in discriminating PD vs. HC; 0.8 (95%CI 0.67, 0.93) for classifying PD vs. OND; and 0.94 (95%CI 0.91, 0.98) for distinguishing PD from all other study participants (i.e. HC+OND). **Table 3** summarizes results for those 163 participants who completed both phases. Score results of the larger, entire cohort (including those had only completed part of the study components) are shown in **Supplementary Table 3**. The latter findings supported the conclusions drawn above.

### PD-RISK Index thresholds for possible clinical decision-making

To examine thresholds, we focused on the comparison of PD vs. HC+OND, which is of greatest relevance to clinical practitioners. Using optimal thresholds corresponding to maximum Youden indices, as shown in **Table 4**, we first visualized in **Supplementary Fig 2** how each score classified participants into two groups, i.e. those ‘with’ vs. those ‘without’ PD. The results demonstrated that the simplified questionnaire and the PD-RISK Index corrected some classification errors made by their original counterparts, which resulted in increased sensitivity for the simplified questionnaire score (from 0.61 to 0.98) and higher specificity for the simplified PD-RISK Index (from 0.7 to 0.83; **Table 4**), respectively. For the comparison between the two smell tests, a trade-off between sensitivity and specificity was observed. Further, when using 22.72 as the threshold (score range, 0–100), the PD-RISK Index of the simplified toolkit identified subjects with PD vs. all other participants in the study at a sensitivity of 0.98, specificity of 0.83, positive likelihood ratio of 5.88, and negative likelihood ratio of 0.02 (**Table 4**).

The Youden index represents one way to determine thresholds; it weighs sensitivity and specificity equally. In practice, other thresholds may be preferred for different clinical scenarios. As a reference for potential clinical decision-making in the future, panels (**B**, **D**, **F**) of **Figs 2**, **3** show various metric values at different thresholds. Besides sensitivity and specificity, the positive likelihood ratio (LR+; calculated as *sensitivity/(1-specificity)*; purple curves) and negative likelihood ratio (LR- ; calculated as *(1-sensitivity)/specificity*; pink curves) represent clinically useful metrics.^29^ Unlike sensitivity and specificity, positive and negative likelihood ratios are not strictly monotonic. As a general rule, using thresholds that are corresponding to higher positive likelihood ratios (e.g. a LR+ >10 signals a strong effect size and LR+ of 5–10 a moderate effect), a positive test result could “rule-in patients with PD”; while using thresholds that are corresponding to lower negative likelihood ratios (e.g. LR- <0.1 for strong, and 0.1–0.2 for moderate effect sizes), a negative test result could effectively “rule-out individuals that do not have PD”. The areas shaded in blue in **Figs 3B**, **3F** represent thresholds for the simplified questionnaire score (range, 129.7–133) and simplified PD-RISK Index (range, 14.78–58.1) that satisfy both a LR+ > 5 and LR- < 0.2; these reflect moderate to high likelihood ranges to “rule-in” and “rule-out” the presence and absence of PD, respectively.^32^ For individual comparisons of metrics for PD vs. HC and PD vs. OND, see **Supplementary Figs 3 and 4**, respectively. (Of note, positive and negative predictive values are also shown for reference in **Figs 2**, **3** and **Supplementary Figs 3, 4**; their values are generally affected by the case–control ratio in a cohort and may thus be less informative than calculating the positive and negative likelihood ratios, which we prioritized here).

**Figure 4.**
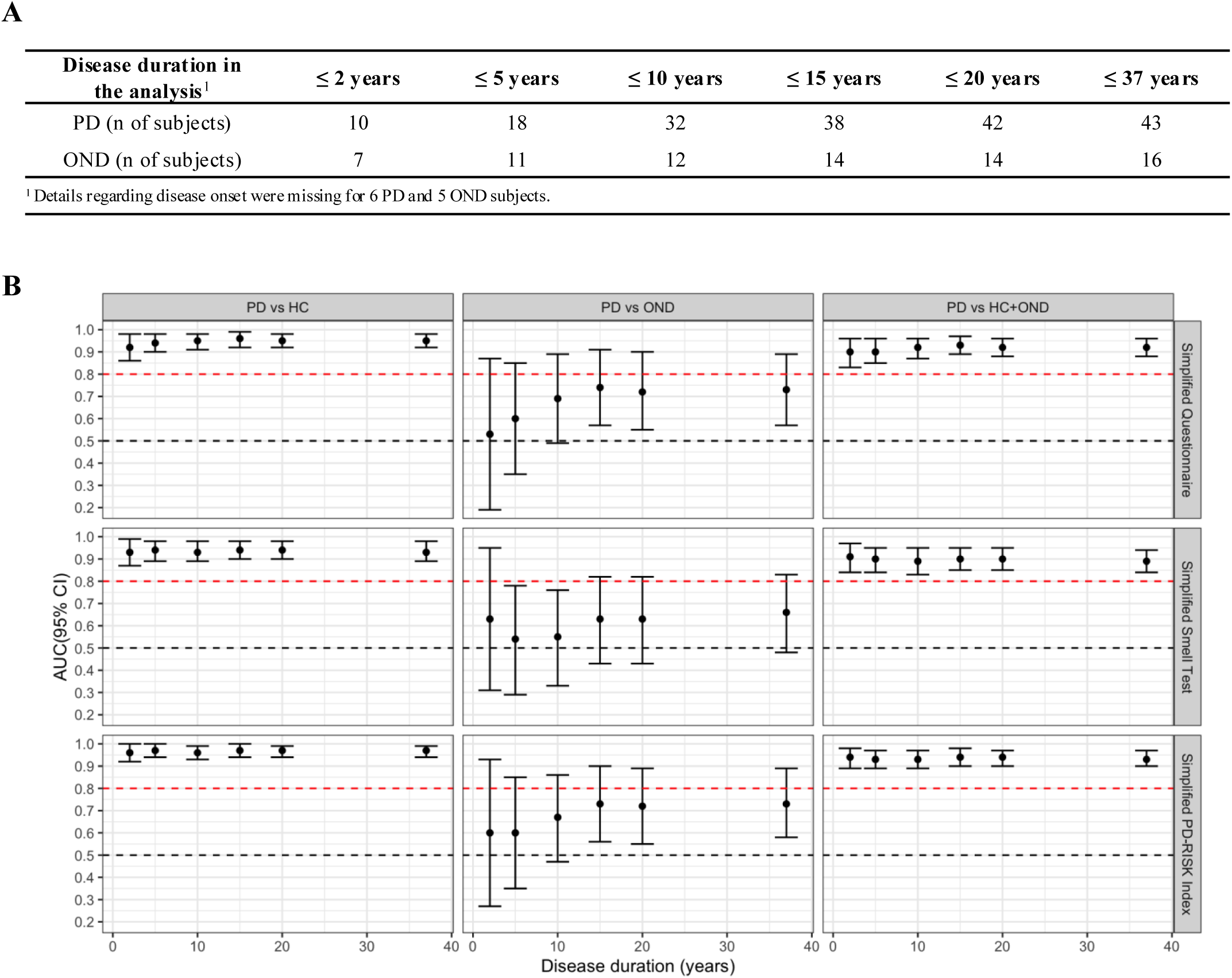
Relationship between duration of disease and discriminative performances for the simplified PD-RISK toolkit. Panel (**A**) lists the number of participants in two neurologically diseased groups when limiting disease duration to the corresponding timeframes in the cohort. Healthy controls remained in these comparisons although disease duration did not apply. In panel (**B**), graphs represent AUC values (y-axes) vs. years since neurological diagnoses were established (x-axis) with error bars representing their 95% confidence interval. Graphs summarize scores of the simplified questionnaire (top row), simplified smell test (middle row), and simplified PD-RISK Index (bottom row), and columns represent diagnostic group comparisons, as indicated. The black and red horizontal, dashed lines represent pre-determined reference levels at 0.5 and 0.8, respectively. PD = Parkinson disease. OND = other neurological diseases. HC = healthy controls. AUC = area under the receiver operating characteristic curve.

### Comparison of score performance between sexes

**Supplementary Table 4** lists effect sizes for each PD-RISK Index variable for both sexes; there, most conclusions remained the same as when both were combined (see **Supplementary Table 1**). In our cohort, the effect of farm life exposure was only significant for females, and a positive history of depressed mood was only significantly associated with higher PD risk in males. **Supplementary Table 5** shows that when examining females vs. males separately, their scores in both versions of the toolkit showed comparable discriminative performances (i.e. AUC values) in their group classifications. The only scenario where a sex bias could be seen was in the comparison of PD vs. OND in the original questionnaire score, which could be partially explained by the smaller sample size and broader range of diseases recruited into the OND group. However, due to the numeric difference by which sex has been incorporated in the questionnaire scores of the PD-RISK Index^11^ (**Table 2**) and due to the known sex effect on olfaction scores (see sex-specific score distributions in **Supplementary Fig 5** and median scores in **Supplementary Table 5**), specific thresholds were determined for each sex; this, to achieve maximum Youden indices and to generate “rule-in” and “rule-out” ranges, as shown in **Supplementary Tables 6**, **7** and **Supplementary Figs 6**, **7**, respectively.

### Index performance vs. time since diagnosis of PD

The simplified version in **Fig 4** and the original version in **Supplementary Fig 8** show AUC values for each score component when examining disease duration from up to 2 years vs. up to 5, to 10, to 15, to 20, and to 37 years. There, the discriminating performance of each score was generally stable regardless of disease duration, especially for components of the simplified PD-RISK Index toolkit, which consistently showed mean AUC values of > 0.9 in correctly classifying PD vs. HC+OND; this finding supports its potential usage in both early and late stages of typical PD.

### Relationship between subjective and objective hyposmia

Finally, we also examined whether female and male participants in each group differed in the assessment of their sense of smell and whether they knew of any explanation for hyposmia when subjects thought it was present (asked in the original PD-RISK Index questionnaire prior to smell testing, see **Supplementary Table 2**). **Supplementary Fig 9** shows that the responses to the question for the subjective presence of any chronic hyposmia-albeit informative-were not as accurate as the results obtained when testing it objectively. Several hyposmic/anosmic participants were unaware of their reduced sense of smell. Other individuals, who had indicated that they experienced hyposmia, showed a normal sense of smell when tested; specifically, several participants, who had indicated chronic olfaction reduction from a known cause (e.g. recurrent or chronic infections, allergy, or injury), had the same range for smell test scores as subjectively normosmic subjects. Interestingly, patients with cognitive impairment (i.e. the MCI/DEM subgroup) were often not aware of their hyposmia, despite very low smell test scores (see orange points in **Supplementary Fig 9**). These findings were independent from what version of smell test was used and of the proband’s sex.

## Discussion

In our study, two versions of the PD-RISK Index toolkit (where the toolkit encompassed a questionnaire, smell test, and an Index score) were evaluated and compared in their discriminative performances to distinguish PD patients from neurologically healthy controls and subjects with other neurological diagnoses. There, we made the following observations: Scores based on questionnaires and smell tests showed high accuracy in the comparison of PD vs. HC groups and moderate performance in the classification of PD vs. OND groups. Versions of the integrated PD-RISK Index scores revealed better discriminative performance than the corresponding scores of individual assessments, indicating that information gathered from known risk factors for PD and for current olfaction performance were synergistic. The latter case was highlighted for example by the comparison of subjective vs. objective olfaction assessment. Further, the simplified version of the toolkit markedly outperformed the original version. Therefore, it will be the preferred tool in our future research efforts and for potential deployment in routine clinic settings and population health studies.

Compared with other, carefully curated and matched case-control studies of PD, ours represents a cohort of participants routinely encountered at outpatient neurology clinics, where subjects were serially invited to participate in a research study. They comprised those with typical PD, and importantly, those with other chronic, neurological conditions, such as atypical forms of parkinsonism, non-PD-type tremors, persons with cognitive impairment, or with restless leg syndrome; hence, they were grouped under the OND category. Of note, sex and disease duration (for PD and OND groups) were not sufficiently matched across disease groups. Therefore, to address any potential influence by sex and disease duration, age- and sex-adjusted analyses were made whenever possible and additional sub-analyses were performed. There, we confirmed that all components of the two versions of the toolkit showed comparable discriminative performances for females and males, i.e. they were not sex biased. Nonetheless, sex-specific thresholds, especially for the questionnaire scores, were suggested (see **Supplementary Tables 5–7** and **Supplementary Figs 5–7**). We also found that performances for the six scores tested were generally stable across various disease durations, indicating the potential usability of the PD-RISK Index toolkit in the diagnosis of both early and advanced stages of PD. Because our study cohort did not include prodromal individuals that have not-yet-converted to manifest disease, the predictive performance of the PD-RISK Index model will be tested in several at-risk-of-PD cohorts in future works.

The optimal thresholds reported in **Table 4** and **Supplementary Tables 6**, **7** corresponded to the maximum Youden indices; however, as mentioned, these may not reflect the best thresholds for clinical decision making. Should a PD-RISK Index-based toolkit be considered for future implementation, clinicians could use an approach as shown for example in **Figs 2**, **3** or **Supplementary Figs 3**, **4**, **6**, **7**; this, to determine other thresholds based on specific clinical needs and testing scenarios. Specifically, the simplified PD-RISK Index, with a wider range that satisfied both LR+ > 5 and LR- < 0.2 (see blue shaded region in **Fig 3F**), offers more flexibility in choosing thresholds, thereby reflecting promising performances in distinct clinical scenarios. Further, should a PD-RISK Index be deployed [together with an app to automatically provide score calculation, prompt interpretation and suggestions to the referring practitioner (see below)], optimal thresholds containing decimals could be used. Alternatively, in the scenario where a toolkit would be implemented using a paper-based form, clinicians might prefer rounded thresholds.

To facilitate future work in larger, multicentric efforts, we see the following 6 benefits of the simplified PD-RISK Index concept: i) Its non-invasive nature and ease of completion for the abbreviated components (questionnaire; smell test); ii) Simple deployment at home (or in the waiting room of healthcare facilities) and its completion in an unsupervised manner; iii) Low production and delivery costs; iv) Acceptable compliance rate due to simplicity and brevity of the testing scheme; v) The option of response entries made online using an app with an automatically calculated summary of scores and a report; and vi) General scalability of the toolkit for routine practice in primary care settings and neurology clinics as well as for community-based screening. Of note, based on informal feedback obtained from our participants the original questionnaire and UPSIT booklets required a mean of ∼50 min to complete, whereas the simplified questionnaire of 11 items and the 8-question smell test card together with online data entry requires a fraction of that time.

Besides assessing score performances, the PD-RISK Index study evaluated the feasibility of using home-based, unsupervised toolkits for screening: with olfaction performance testing, 79% of mailed UPSIT booklets and 79.8% of mailed simplified smell test cards were completed, indicating a relatively high compliance rate. Regarding data quality, no major issues were observed for data collected online (including all the questions and entries for the abbreviated smell test responses), and no study participant reported technical difficulties. For data entry using UPSIT, select issues were identified, because: i) Its booklets require test-takers to make a choice even when no odorant could be confidently identified, whereby participants either marked multiple options for one scent, or left some questions unanswered, or simply guessed; and ii) Manually inputting 40 responses for each test submitted requires a research study assistant, and it is more time consuming (which in theory could also be a source of error).

The simplified smell test (NeuroScent^®^ Card, **Supplementary Fig 1**) was designed to optimize data quality and test-taker experience, as gleaned from phase I of the study and participants’ informal feedback. As demonstrated in phase II, the abbreviated test enabled online data-entry by participants themselves. With a 5^th^*“I cannot identify it”* option, participants, especially those with more severe olfaction impairment, could respond more accurately rather than randomly guess or leave a blank response. The current version of the 8-scent card represents a prototype that is undergoing further evaluation at additional sites; it has been deployed in four languages (English, French, German, Spanish), and is currently being further tested in head-to-head comparisons of its performance vs. UPSIT and the Sniffin’ Sticks Identification Test.^33^

Our study has limitations. Due to our approach in recruiting the HC group, there was a known selection bias where neurologically healthy individuals that participated in the study were more likely to have a relationship with or relation to a person with PD, or to have a greater interest in the disease itself, when compared to subjects from the general population; hence, the true effect size of-for example-a positive family history of PD may have been misrepresented in the PD vs. HC groups. Further, OND represents an umbrella term containing many neurological conditions, with rather small sample sizes for each specific disease in this study. Therefore, the results reported here are mostly informative in identifying typical PD patients, but less so for disease differentiation within the group of subjects categorized under OND. Future studies will include more participants with a range of chronic conditions affecting movements and gait in adults that are seen routinely by medical practitioners and neurologists.

Despite our effort to recruit participants from varied race/ethnicity groups, the Ottawa-based PD-RISK Index study cohort encompassed >85% Whites (see **Supplementary Table 2**). Although underpowered, we nonetheless examined the score distribution vs. self-identified race/ethnicity in each assessment. There, we observed no significant effect of the variable race/ethnicity (see **Supplementary Fig 10**). Importantly, because risk for Parkinson’s has been reported to vary across race/ethnic groups^34–39^ and because there is evidence that scent detection and discrimination may vary by race/ethnicity,^40–42^ future work using targeted recruitment techniques and optimized odorant selection will be necessary to test the extent of generalizability of the simplified version used in the current toolkit. Of note, ethnicity related to the question of “Hispanic or Latino (Spanish origin)” and select options pertaining to race (e.g. “Asian”, “Native Hawaiian or other Pacific Islander”, “White”, and “Other (specify which)”) were adopted from the demographic questionnaires used in the original versions of the Harvard PD Biomarker Study cohort^43,44^ and Parkinson’s Progression Marker Initiative (PPMI) study.^45^ Under ‘race’, we added the option “Indigenous Peoples in Canada (the First Nations, Inuit and Métis)” to reflect the population of Canada; options for “Ashkenazi Jew”,^36,37^ “Basque”,^38^ and “African Berber”^39^ were added because of known genetic risk associations between PD and these groups (note, no participant self-identified as “Basque” and “African Berber” here). However, we recognize that some of these options are outdated or unclear, and these questions will be updated with more clarity and description in future versions.

Further, The PD-RISK Index study cohort was used to externally validate both versions of the questionnaire and UPSIT; for some scores, such as those based on the simplified smell test, early results from our study had been used in its creation. Therefore, the role of the cohort and the risk of potential overfitting were described and managed, and future validation studies using external cohorts from other sites are underway. In parallel efforts to further validate the simplified version of the toolkit, to potentially improve it, and to prepare it for wider clinical deployment, we have developed a web-based app (PD-RISK Index calculator); for future probands that enter their responses online, the app calculates scores to automatically generate a report. In the coming months, this simplified toolkit and a prototype app will be validated within the Canadian Open Parkinson Network (C-OPN) cohort^46^ with its >2,500 active participants, including patients with typical PD and atypical parkinsonism, and neurologically healthy controls.

From results shown here, the PD-RISK Index toolkit, especially its simplified version, revealed high accuracy in differentiating PD patients from neurologically healthy controls. For the comparison between PD vs. those with other neurological diagnoses, the toolkit’s performance requires further optimization. We are currently working on enhancing the model’s ability in disease classification of PD vs. OND, such as by adding other, easy-to-collect variables and non-invasive biomarkers, e.g. assessments for orthostatic hypotension^47^ and altered heart rate variability^48^, to the current model. These two indices have been reported as altered in synucleinopathy disorders (i.e. PD; DLB; MSA). Future studies will also explore a potentially better distinction between the MCI/dementia and typical PD groups, such as by adding questions that sensitively probe for early cognitive difficulties and/or behavioural changes (e.g. anosognosia, with the inability to recognize or communicate the presence of one’s hyposmia). With further improvements to the current toolkit and appropriate validation steps at locations elsewhere, we envision a future when non-neurologists will be able to confidently identify the high likelihood of PD being present in an adult, and possibly, of a prodromal state of the syndrome; a scenario that should be based on scientific evidence and a non-invasive, inexpensive and simple-to-deploy assessment tool with rapid feedback, such as of a metric value (‘Index’).

In conclusion, the PD-RISK Index model represents a simplified, home-based method with relatively high sensitivity and specificity to identify PD patients. As such, it offers a potentially scalable solution to bridge the delay between a general practitioner’s assessment and specialty consultation, thereby facilitating earlier symptomatic treatment with already approved therapeutics and physiotherapy.^3^ Finally, the toolkit may also enable effective screening of and risk quantification in individuals that seek enrolment into primary prevention studies; this, to measurably lower the likelihood of developing manifest PD in the future or delay its onset.^49^ Such initiatives are currently pursued by multiple organizations worldwide.^10,45,50^

## Acknowledgements

The authors acknowledge the commitment of study participants in the Ottawa PD-RISK Index study. We thank Nadine Mauri, Nancy MacDonald, and Yoobin Lee for help in data management and input. We are grateful for the ongoing support and feedback from people with lived experiences, such as through the board of the ‘Parkinson Research Consortium’ and members of the ‘Partners Investing in Parkinson’s Research’ group (both in Ottawa), for their ongoing encouragement.

## Author contributions

MGS and JL developed the concept and design of the study, DL and NS contributed ideas to the questionnaire design; KG, JS, AF, MGS, JJT and JL contributed to data collection and verification. JL and DM decided on the statistical methods used in this study. JL performed data cleaning, data analysis, figures and tables. All authors contributed to data interpretation. JL and MGS wrote the first draft of the manuscript. All authors contributed to the editing of the manuscript and revising it critically, and all authors approved the submission of its current version.

## Statements and declarations

### Ethical considerations

The study was approved by the Ottawa Health Science Network Research Ethics Board at TOH (REB #: 20210308-01H) and the Research Ethics Board at EBH (REB #: M16-15-050).

### Consent to participate

As approved by the Research Ethics Boards at both TOH and EBH, participants consented via *Implied Consent*, with the following language highlighted in the two online questionnaires: “*CONSENT: By completing this questionnaire your consent to participate is implied.*”

### Consent for publication

Only aggregated results are published in the manuscript. Carefully de-identified data will be made publicly available (see “Data availability”). Information regarding data sharing was included in the “Implied Consent Form”.

## Data availability

This study used de-identified data of the PD-RISK Index study cohort. Data will be further de-identified and made publicly available (link to be updated, further deidentification process is required). If needed, please contact the authors for access to the complete original data.

## Code availability

The code for data analyses, figures and tables is publicly accessible in GitHub https://github.com/JuanLiOHRI/PDRI.

## aSCENT-PD Investigators

Ben Arenkiel^5,11^, Zhandong Liu^5,11^, Brit Mollenhauer^5,12,13,14^, Josef Penninger^5,15^, Max Rousseaux^5,16^, Armen Saghatelyan^5,16^, Natalina Salmaso^5,10^, Michael G. Schlossmacher^1,4,5,6*^, Christine Stadelmann^5,17^, Julianna J. Tomlinson^1,4,5^, John M. Woulfe^1,5^

^11^ Baylor College of Medicine, Houston, TX, United States

^12^ Paracelsus-Elena-Klinik, Kassel, Germany

^13^ Department of Neurology, University Medical Center Goettingen, Goettingen, Germany

^14^ Deutsches Zentrum für Neurodegenerative Erkrankungen (DZNE), Goettingen, Germany

^15^ The University of British Columbia, Vancouver, BC, Canada

^16^ Department of Cellular and Molecular Medicine, University of Ottawa, Ottawa, ON, Canada

^17^ Institute for Neuropathology, University Medical Center Goettingen, Goettingen, Germany

---

**Supplementary Figure 1.**
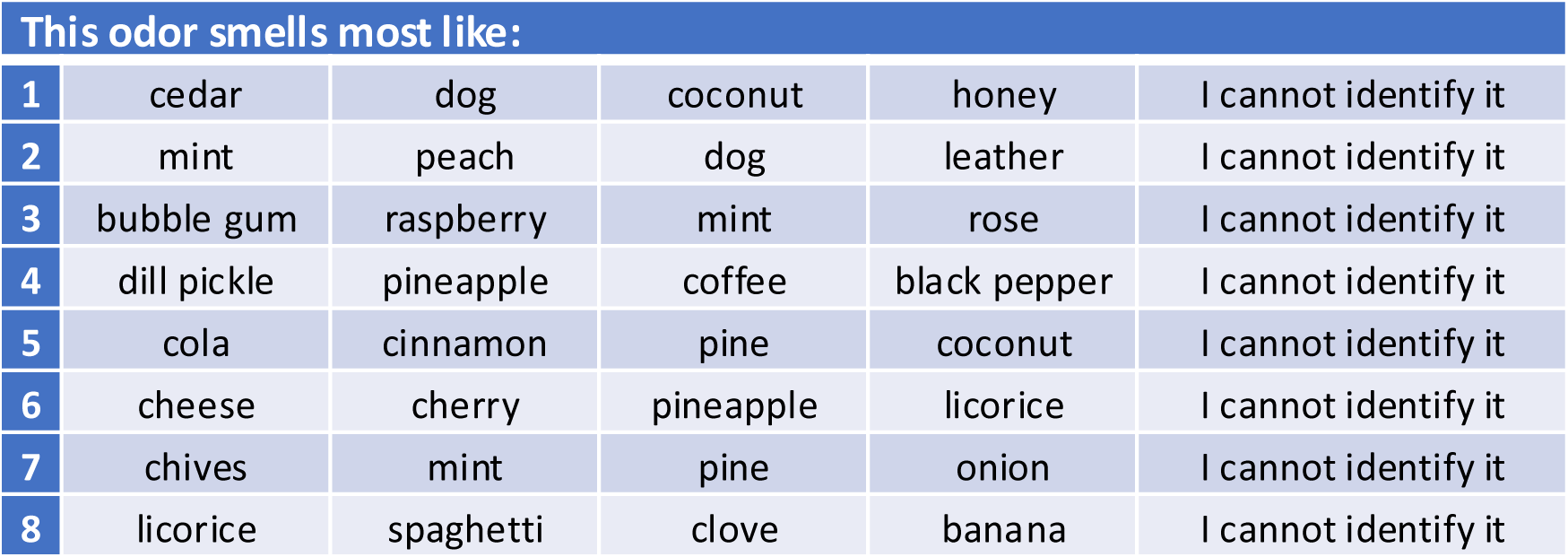
The simplified smell test. Participants were asked to identify seven odorants: dill pickle, mint (twice), rose, coconut, cinnamon, clove, and licorice.

**Supplementary Figure 2.**
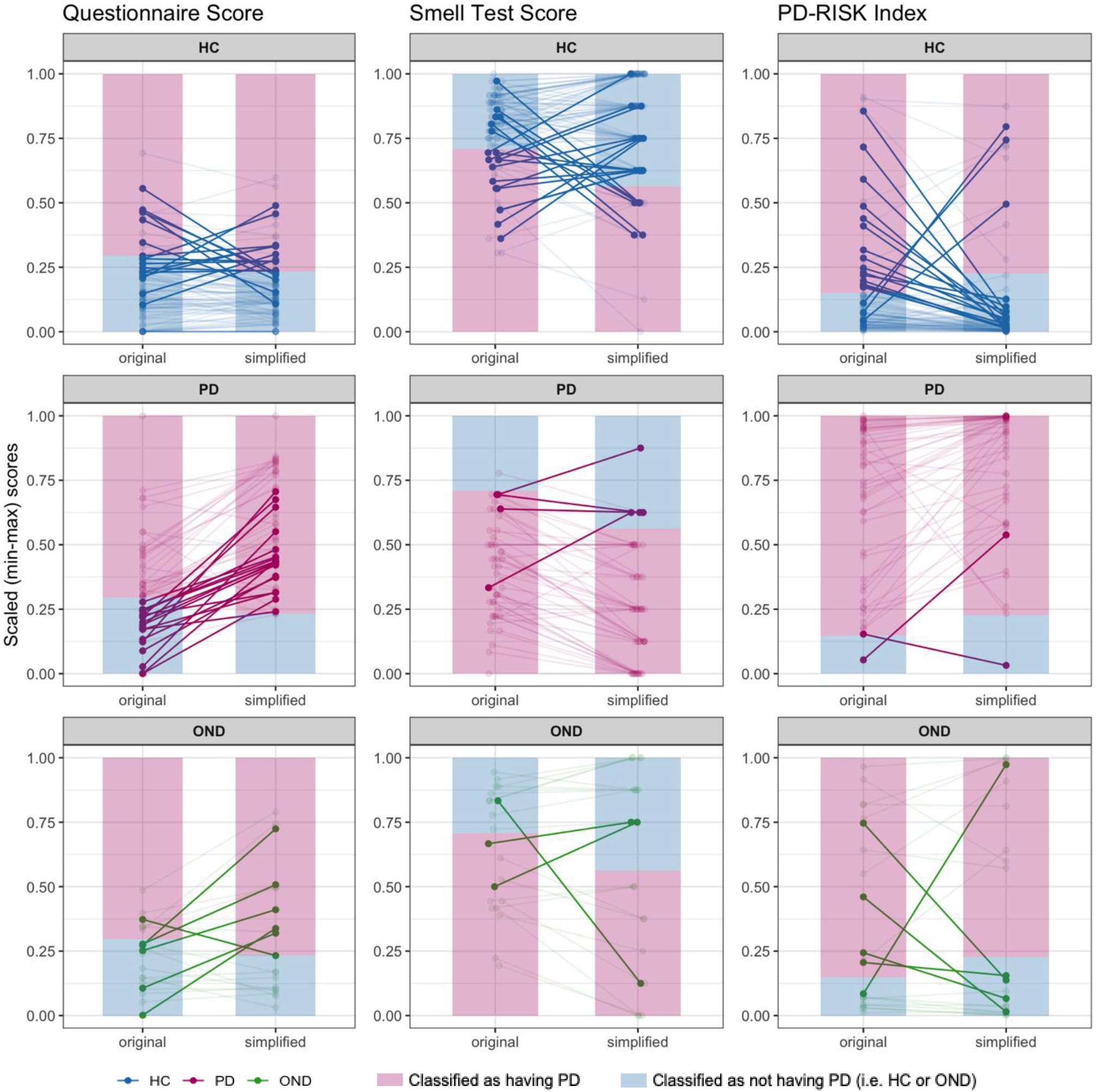
Visualization of classification and reclassification for scores in the *original vs. simplified version* of the PD-RISK toolkit, respectively. The left, middle and right columns represent versions of the questionnaire, smell test, and PD-RISK Index, respectively. The top, middle, bottom rows represent classifications of HC, PD, OND subjects, respectively. Each data point represents the score of one participant, and lines connect the pair of scores from the original to the simplified version for each participant; colors represent participants’ actual diagnostic group, as shown in the legend below. To better visualize comparisons, scores were rescaled to range [y-axes; from 0 to 1] by min-max scaling. Vertical areas shaded in light pink and light blue represent the corresponding decision areas as classified by scores as “having PD” or “not having PD”, respectively, using the corresponding optimal thresholds for PD vs. HC+OND [(rescaled); see **Table 4**]. Study participants whose group classification did not match between the two versions of the toolkit (and were re-classified) are highlighted by solid lines and points. HC = healthy controls. PD = Parkinson disease. OND = other neurological diseases.

**Supplementary Figure 3.**
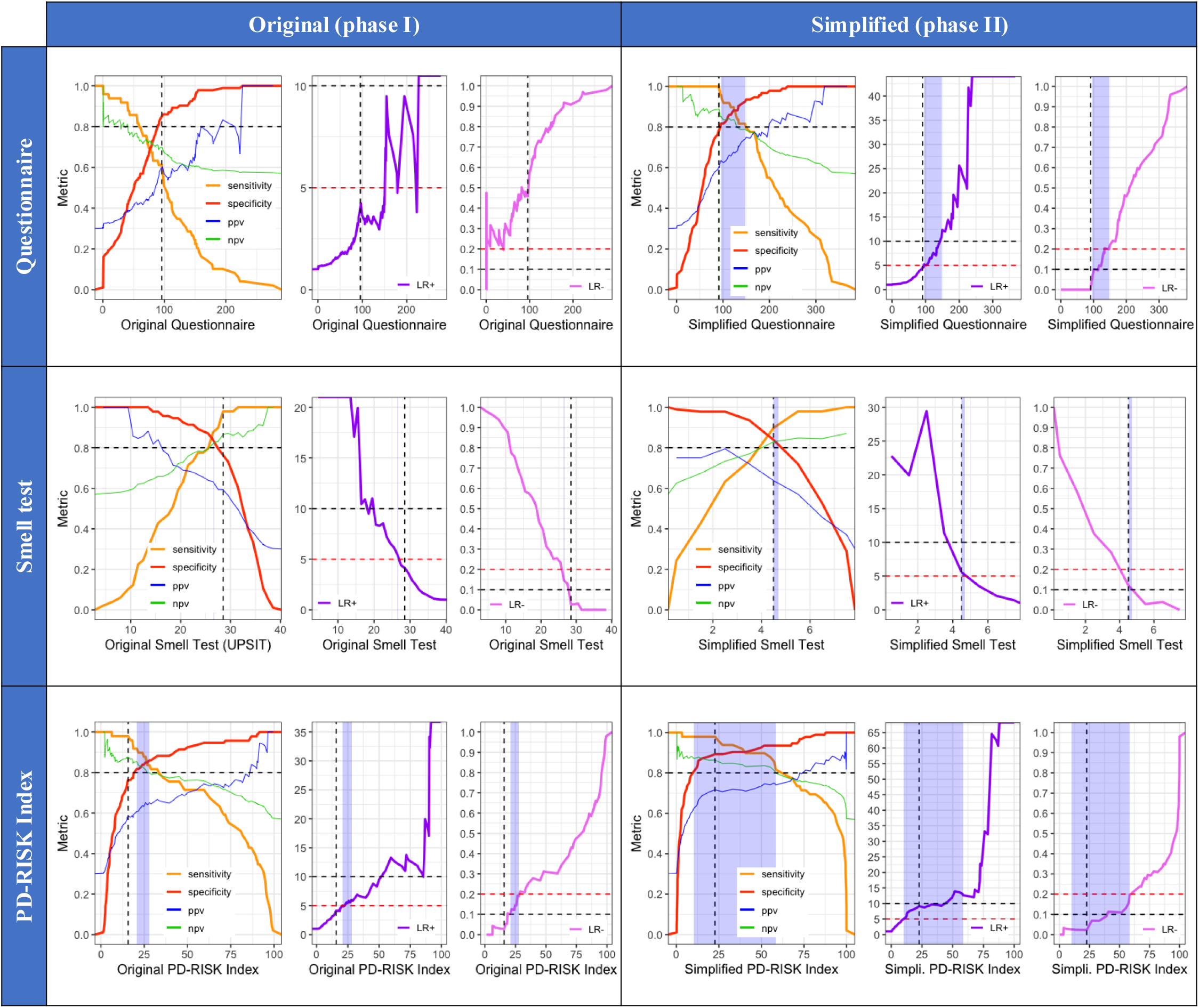
Discriminative performances of two versions in the PD-RISK toolkit for *comparing PD vs. HC* groups in the Ottawa-based study. The left and right columns represent the original toolkit in phase I and the simplified toolkit in phase II, respectively. The top, middle and bottom rows represent versions of the questionnaire, smell test, and PD-RISK Index, respectively. Colors represent different metrics, as shown in the legend. The vertical dashed line in each figure represents the corresponding optimal threshold as shown in **Table 4**. Horizontal dashed lines represent various reference values: sensitivity, specificity, PPV, and NPV values at 0.8; LR+ values at 5 (red) and 10 (black); and LR- values at 0.2 (red) and 0.1 (black). The shaded (light blue) areas represent thresholds that satisfy both LR+ > 5 and LR- < 0.2. PD = Parkinson disease. HC = healthy controls. PPV = positive predictive value. NPV = negative predictive value. LR+ = positive likelihood ratio. LR- = negative likelihood ratio. UPSIT = University of Pennsylvania Smell Identification Test.

**Supplementary Figure 4.**
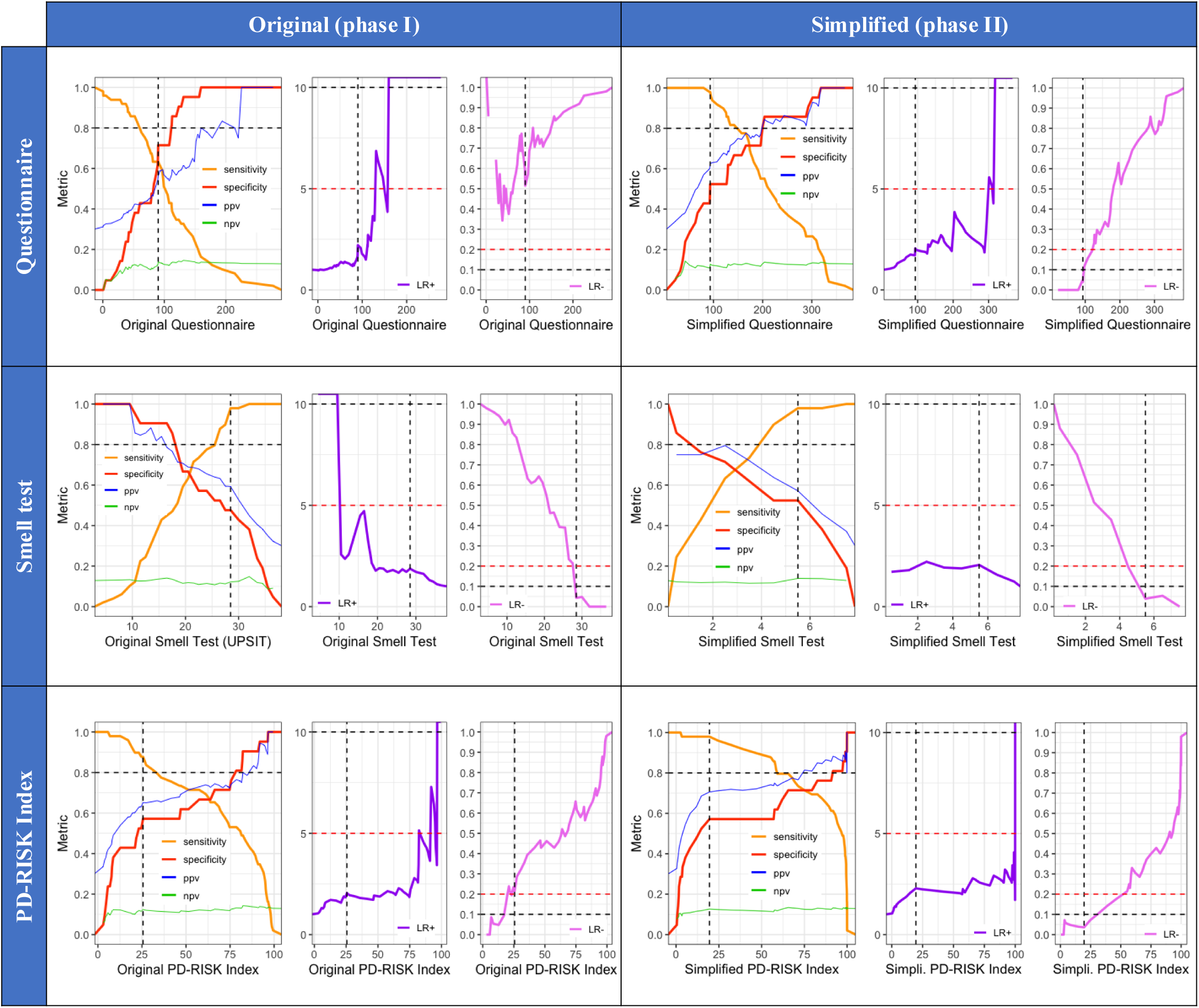
Discriminative performances of two versions in the PD-RISK toolkit for *comparing PD vs. OND* groups in the Ottawa-based study. The left and right columns represent the original toolkit in phase I and the simplified toolkit in phase II, respectively. The top, middle and bottom rows represent versions of the questionnaire, smell test, and PD-RISK Index, respectively. Colors represent different metrics, as shown in the legend. The vertical dashed line in each figure represents the corresponding optimal threshold as shown in **Table 4**. Horizontal dashed lines represent various reference values: sensitivity, specificity, PPV, and NPV values at 0.8; LR+ values at 5 (red) and 10 (black); and LR- values at 0.2 (red) and 0.1 (black). PD = Parkinson disease. OND = other neurological diseases. PPV = positive predictive value. NPV = negative predictive value. LR+ = positive likelihood ratio. LR- = negative likelihood ratio. UPSIT = University of Pennsylvania Smell Identification Test.

**Supplementary Figure 5.**
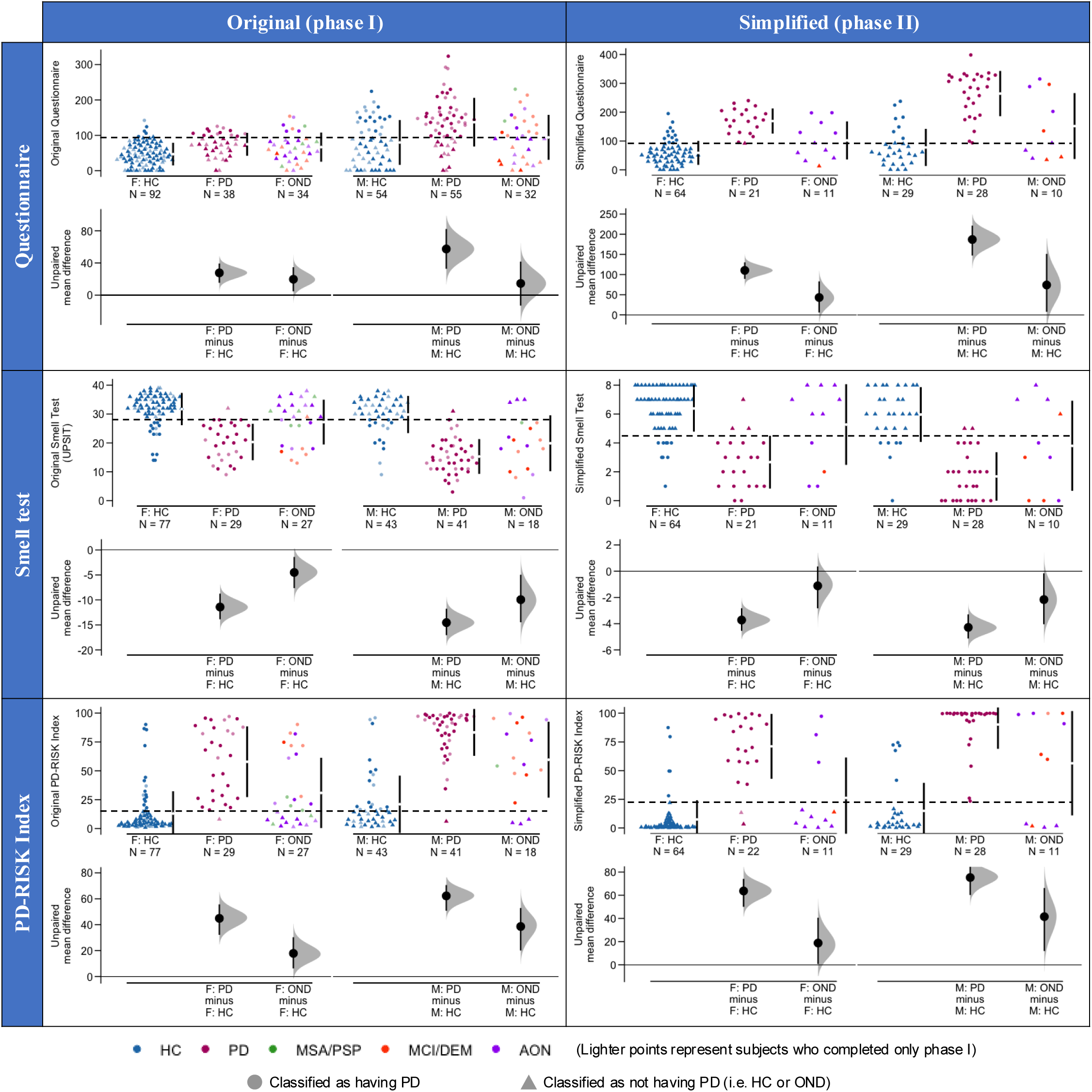
Distribution of scores using two versions of the PD-RISK toolkit for different diagnostic groups in the Ottawa-based cohort, as *separated by sex*. Cummings estimation plots were used to illustrate and compare score distributions in each diagnostic group and sex: the left and right columns are for the original toolkit in phase I and simplified toolkit in phase II, respectively. The top, middle, bottom rows represent versions of the questionnaire, smell test, and PD-RISK Index, respectively. Each data point in the upper panel of each graph represents the score of one participant, and colors represent different groups and diagnoses, as shown in legends. In left panels, data points in lighter colors represent participants who only completed tests in the phase I of the trial. The horizontal dashed lines in the upper panels of each graph represent the corresponding optimal thresholds reported in **Table 4** for PD vs HC + OND, when both sexes combined; participants were represented by circular data points when classified as having PD; otherwise, they were represented by data points as triangles (see legend). Vertical lines in the upper panel of each graph represent the conventional mean ± standard deviation error bars. Lower panels show mean group differences (for effect size) and 95% confidence intervals, as estimated by bias-corrected and accelerated bootstrap, using healthy controls within each sex as the reference group. F = Female. M = Male. UPSIT = University of Pennsylvania Smell Identification Test. HC = healthy controls. PD = Parkinson disease. DLB = dementia with Lewy bodies. OND = other neurological diseases. MSA = multiple system atrophy. PSP = progressive supranuclear palsy. MCI = mild cognitive impairment. DEM = dementia. AON = all other neurological diseases.

**Supplementary Figure 6.**
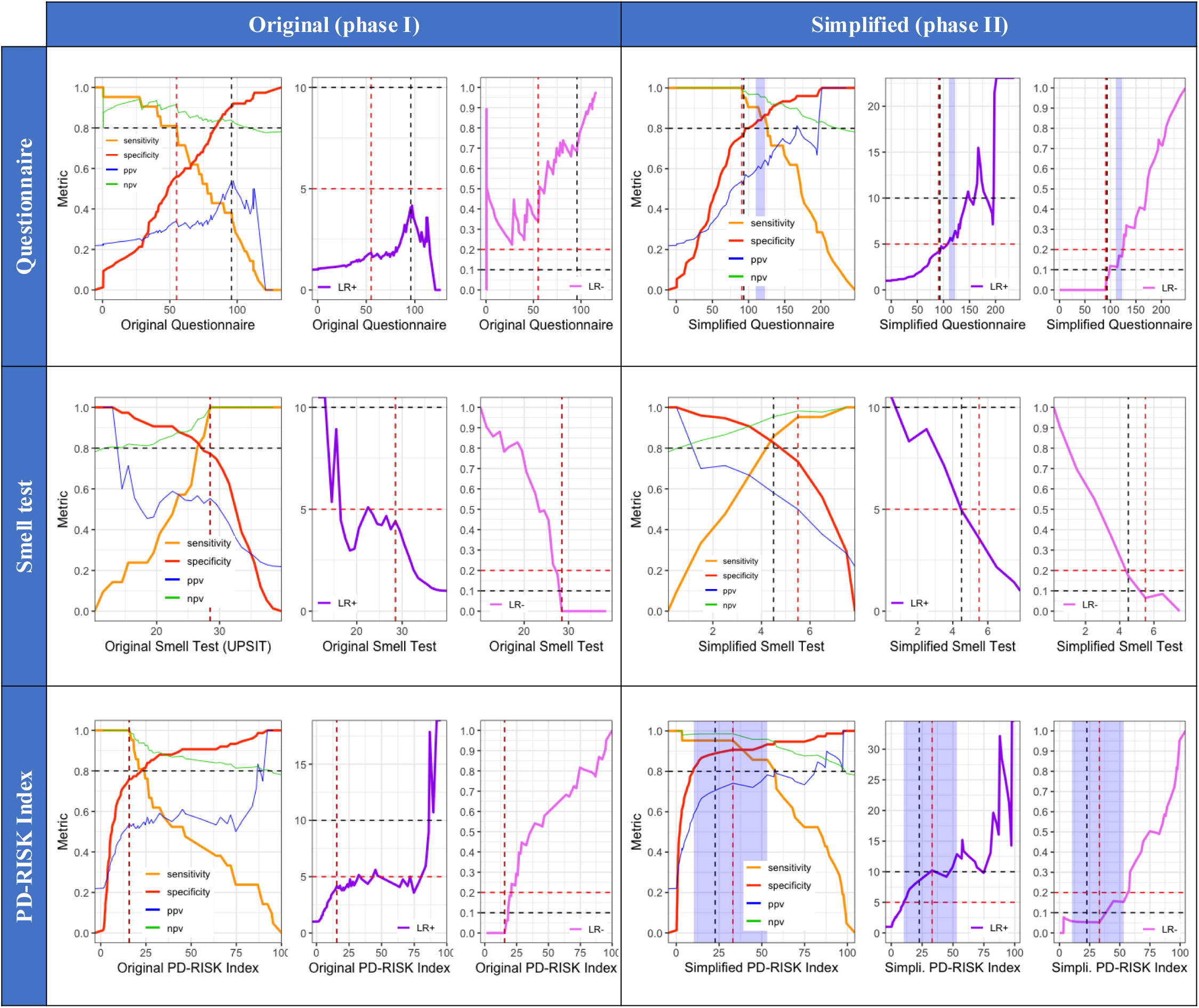
Discriminative performances of two versions of the PD-RISK toolkit for comparing PD vs. HC+OND groups in the Ottawa-based trial, shown for *female* participants. The left and right columns represent the original toolkit in phase I and the simplified toolkit in phase II, respectively. The top, middle and bottom rows represent versions of the questionnaire, smell test, and PD-RISK Index, respectively. Colors represent different metrics, as shown in the legend. The black and red vertical dashed line in each figure represents the corresponding sex-combined and sex-specific optimal threshold, respectively, as shown in **Supplementary Table 6**. Horizontal dashed lines represent various reference values: sensitivity, specificity, PPV, and NPV values at 0.8; LR+ values at 5 (red) and 10 (black); and LR- values at 0.2 (red) and 0.1 (black). The shaded areas (light blue) represent thresholds that satisfy both LR+ > 5 and LR- < 0.2. PD = Parkinson disease. HC = healthy controls. OND = other neurological diseases. PPV = positive predictive value. NPV = negative predictive value. LR+ = positive likelihood ratio. LR- = negative likelihood ratio. UPSIT = University of Pennsylvania Smell Identification Test.

**Supplementary Figure 7.**
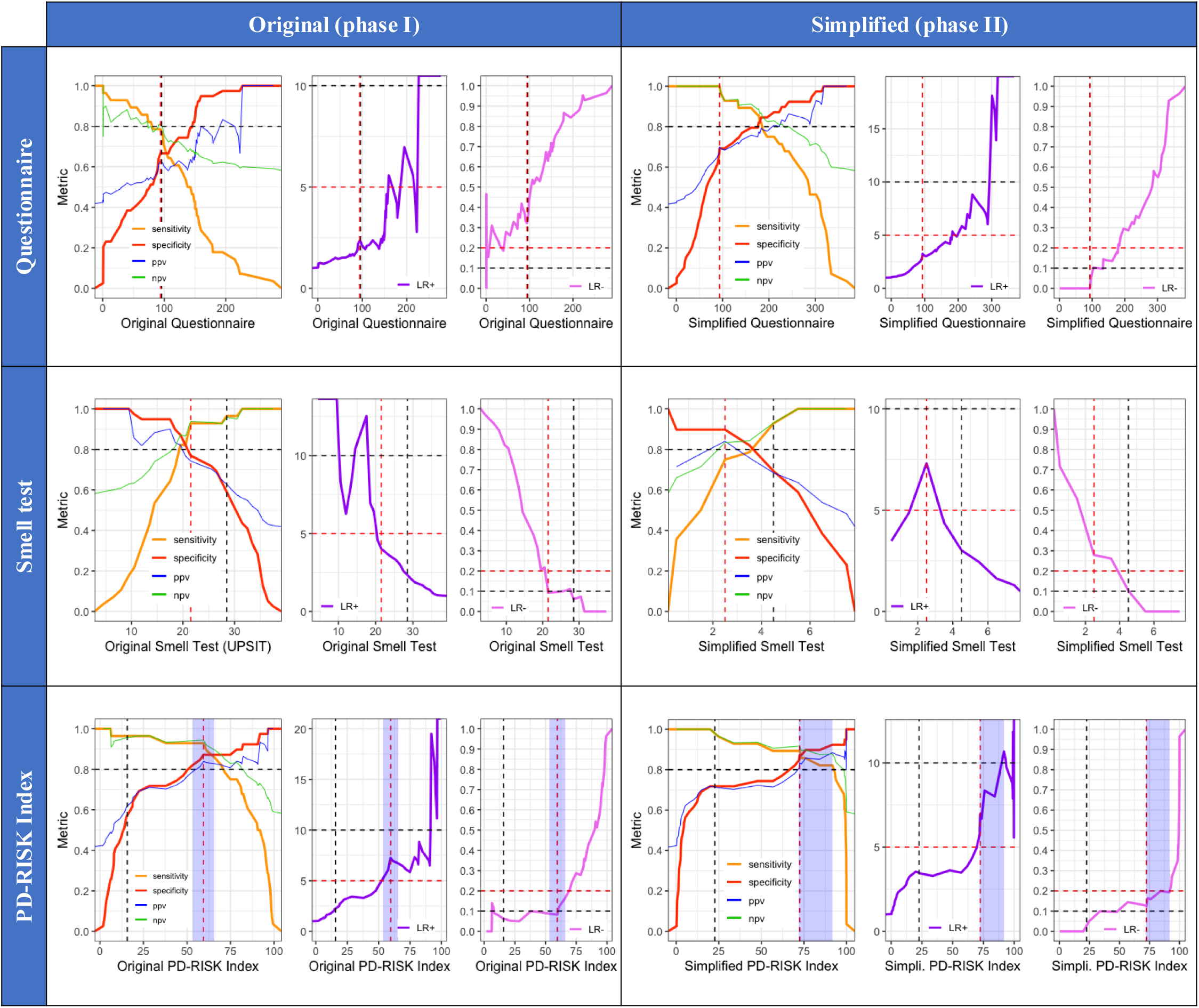
Discriminative performances of two versions of the PD-RISK toolkit for comparing PD vs. HC+OND groups in the Ottawa-based trial, shown for *male* participants. The left and right columns represent the original toolkit in phase I and the simplified toolkit in phase II, respectively. The top, middle and bottom rows represent versions of the questionnaire, smell test, and PD-RISK Index, respectively. Colors represent different metrics, as shown in the legend. The black and red vertical dashed line in each figure represents the corresponding sex-combined and sex-specific optimal threshold, respectively, as shown in **Supplementary Table 7**. Horizontal dashed lines represent various reference values: sensitivity, specificity, PPV, and NPV values at 0.8; LR+ values at 5 (red) and 10 (black); and LR- values at 0.2 (red) and 0.1 (black). The shaded areas (light blue) represent thresholds that satisfy both LR+ > 5 and LR- < 0.2. PD = Parkinson disease. HC = healthy controls. OND = other neurological diseases. PPV = positive predictive value. NPV = negative predictive value. LR+ = positive likelihood ratio. LR- = negative likelihood ratio. UPSIT = University of Pennsylvania Smell Identification Test.

**Supplementary Figure 8.**
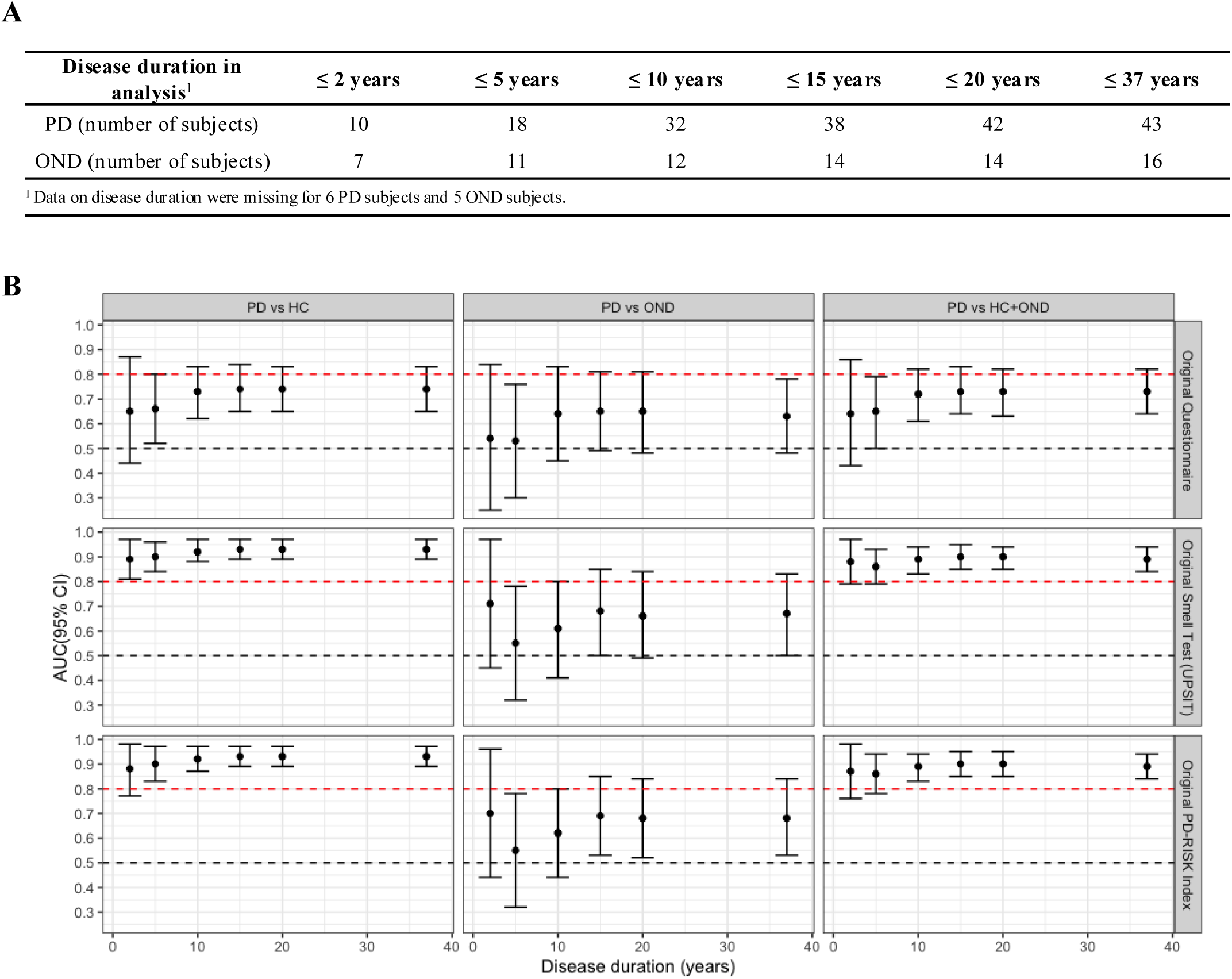
Relationship between length of disease and discriminative performances for the *original* PD-RISK toolkit. Panel (**A**) lists the number of participants in two neurologically diseased groups when limiting disease duration to the corresponding timeframes in the cohort. Healthy controls remained in these comparisons although disease duration did not apply. In panel (**B**), graphs represent AUC values (y-axes) vs. years since neurological diagnoses were established (x-axis) with error bars representing their 95% confidence interval. Graphs summarize scores of the simplified questionnaire (top row), simplified smell test (middle row), and simplified PD-RISK Index (bottom row), and columns represent diagnostic group comparisons, as indicated. The black and red horizontal, dashed lines represent pre-determined reference levels at 0.5 and 0.8, respectively. PD = Parkinson disease. OND = other neurological diseases. HC = healthy controls. AUC = area under the receiver operating characteristic curve. UPSIT = University of Pennsylvania Smell Identification Test.

**Supplementary Figure 9.**
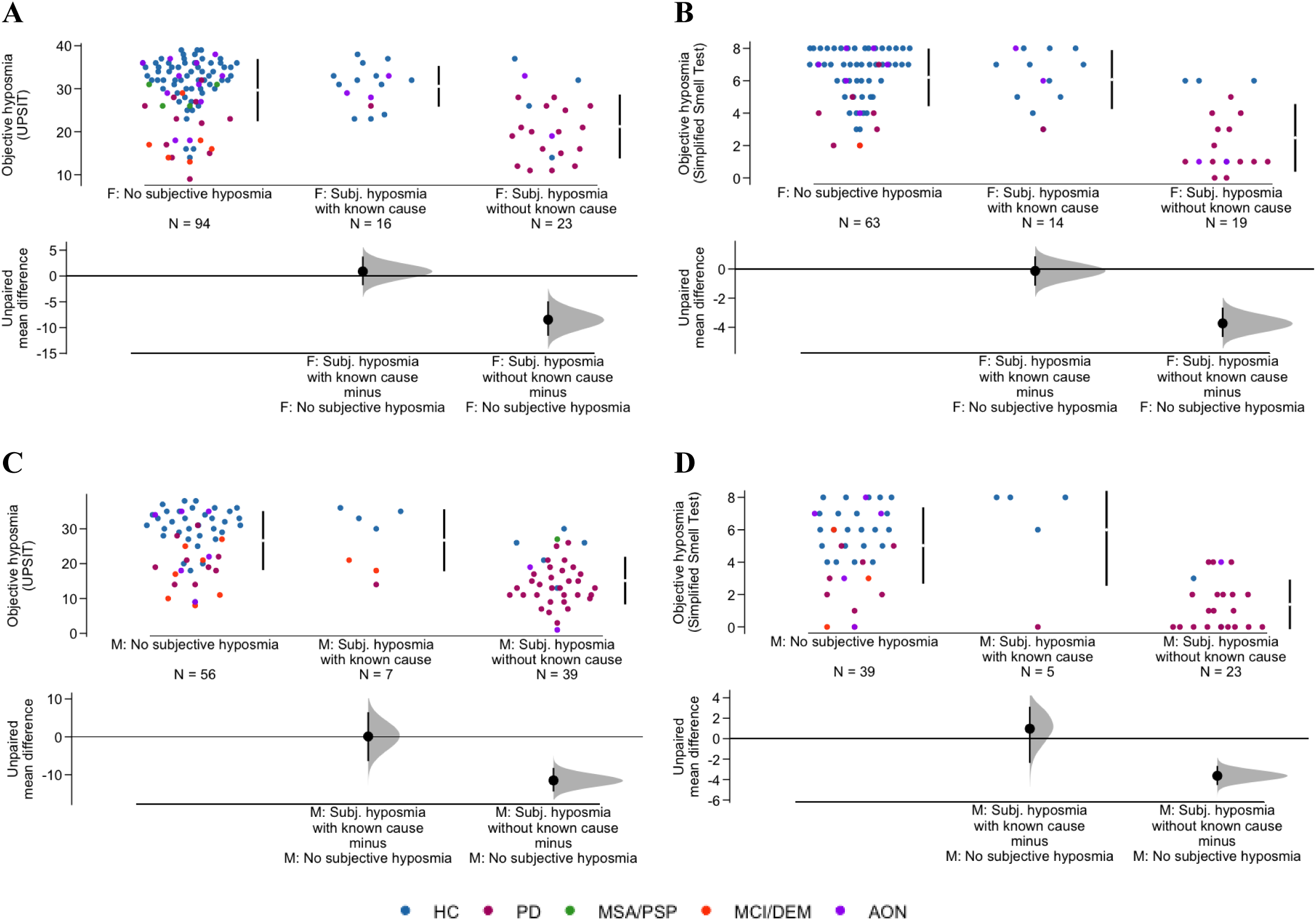
Relationship between subjective report vs. objective assessment of chronic hyposmia, as captured by two smell test versions, as *separated by sex*. Cummings estimation plots were used to illustrate score distributions in each group of persons regarding subjective hyposmia. The left (**A**, **C**) and right (**B**, **D**) columns represent scores of UPSIT and the simplified smell test, respectively. The upper (**A**, **B**) and lower (**C**, **D**) rows are for female and male participants, respectively. Each data point in the upper panel of each figure represents the score of one participant, and colors represent different groups and diagnoses, as shown in the legend; for exact diagnosis, see **Table 1**. Vertical lines in the upper panel of each graph represent the conventional mean ± standard deviation error bars. Lower panels show the mean group difference (*i.e.*, effect size) and its 95% confidence interval estimated by bias-corrected and accelerated bootstrapping, using participants who did not report subjective hyposmia as the reference group. Known causes of subjective olfaction reduction included: COVID-19, seasonal allergy / allergic rhinitis, recurrent nasal cavity / sinus infection, and facial / nasal / skull fracture. F = Females. M = Males. UPSIT = University of Pennsylvania Smell Identification Test. HC = healthy controls. PD = Parkinson disease. OND = other neurological diseases. MSA = multiple system atrophy. PSP = progressive supranuclear palsy. MCI = mild cognitive impairment. DEM = dementia. AON = all other neurological diseases.

**Supplementary Figure 10.**
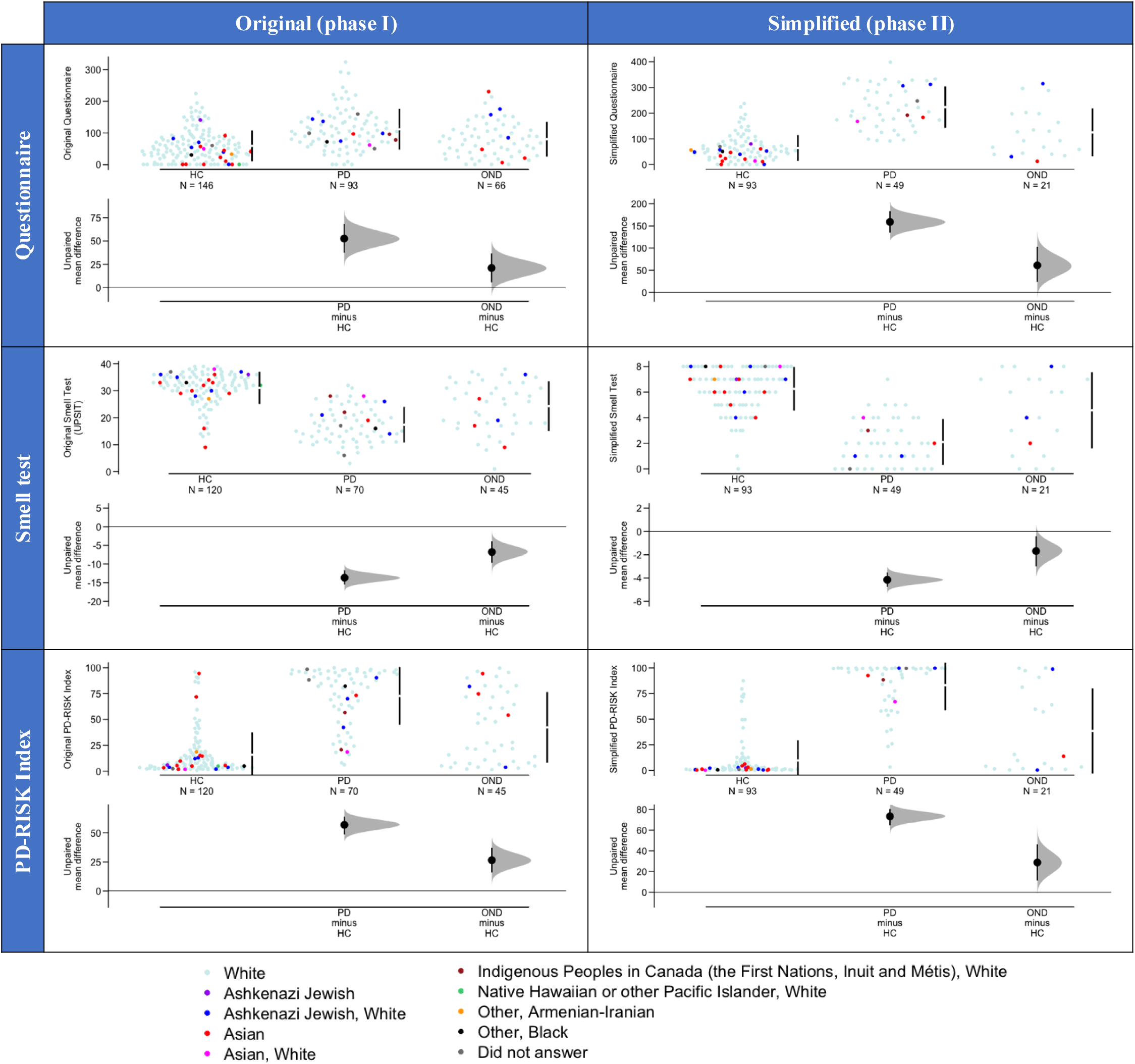
Distribution of scores using two versions of the PD-RISK toolkit for different diagnostic groups, with *self-identified race/ethnicity.* Cummings estimation plots were used to illustrate and compare score distributions in each diagnostic group: the left and right columns are for the original toolkit in phase I and simplified toolkit in phase II, respectively. The top, middle, bottom rows represent versions of the questionnaire, smell test, and PD-RISK Index, respectively. Each data point in the upper panel of each graph represents the score of one participant, and colors represent self-identified races (see legend). Vertical lines in the upper panel of each graph represent the conventional mean ± standard deviation error bars. Lower panels show mean group differences (for effect size) and 95% confidence intervals (CI), as estimated by bias-corrected and accelerated bootstrap, using healthy controls as the reference group. HC = healthy controls. PD = Parkinson disease. OND = other neurological diseases. UPSIT = University of Pennsylvania Smell Identification Test.

## SUPPLEMENTARY TABLES

**Supplementary Table 1:**
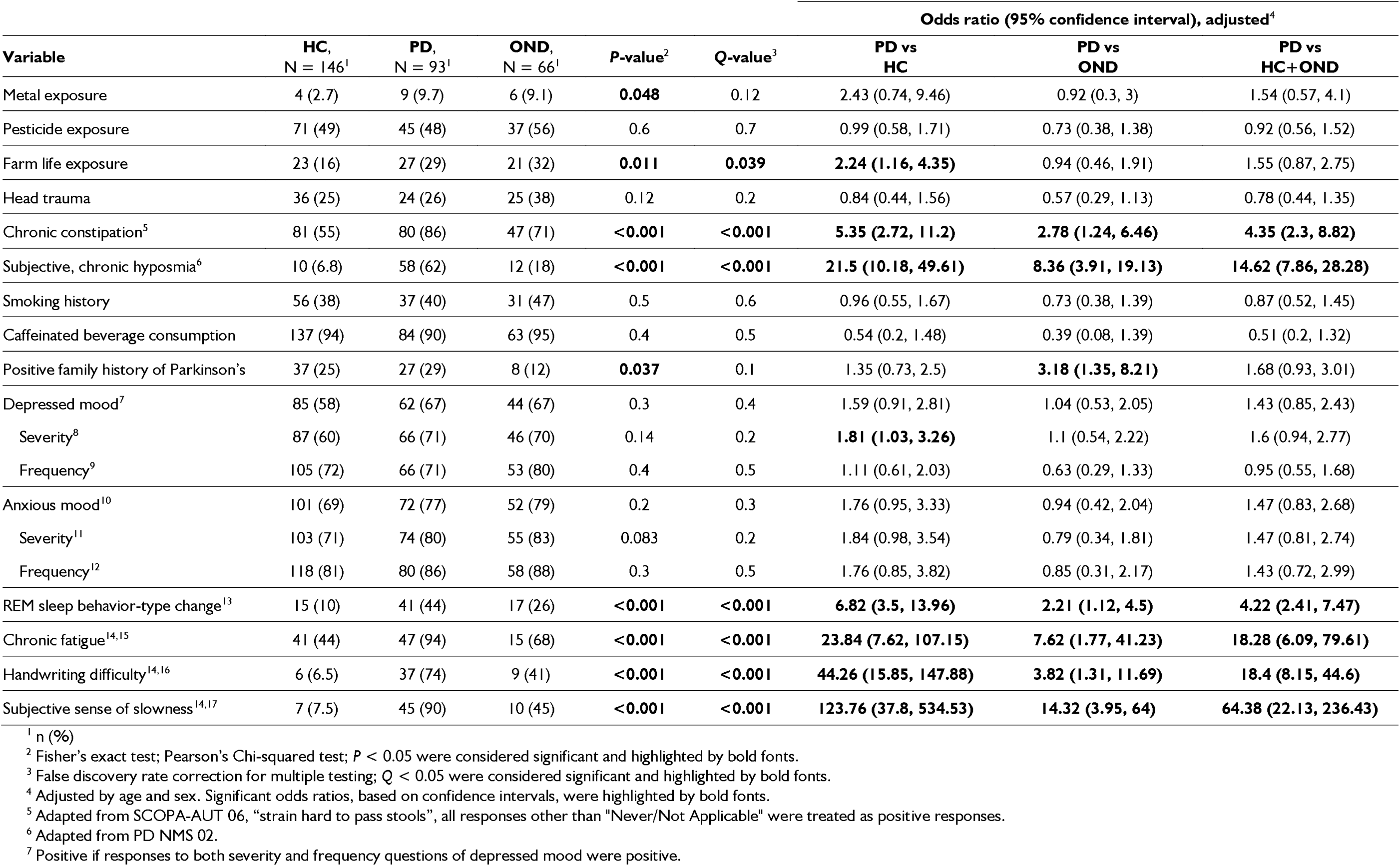
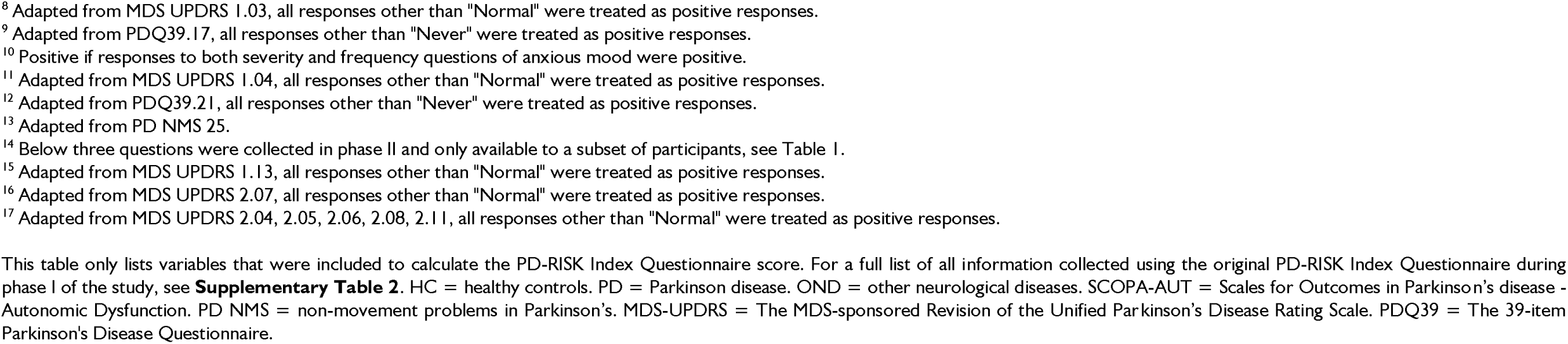
Summary of odds ratios for variables collected in the two versions of the PD-RISK Index questionnaire.

**Supplementary Table 2:**
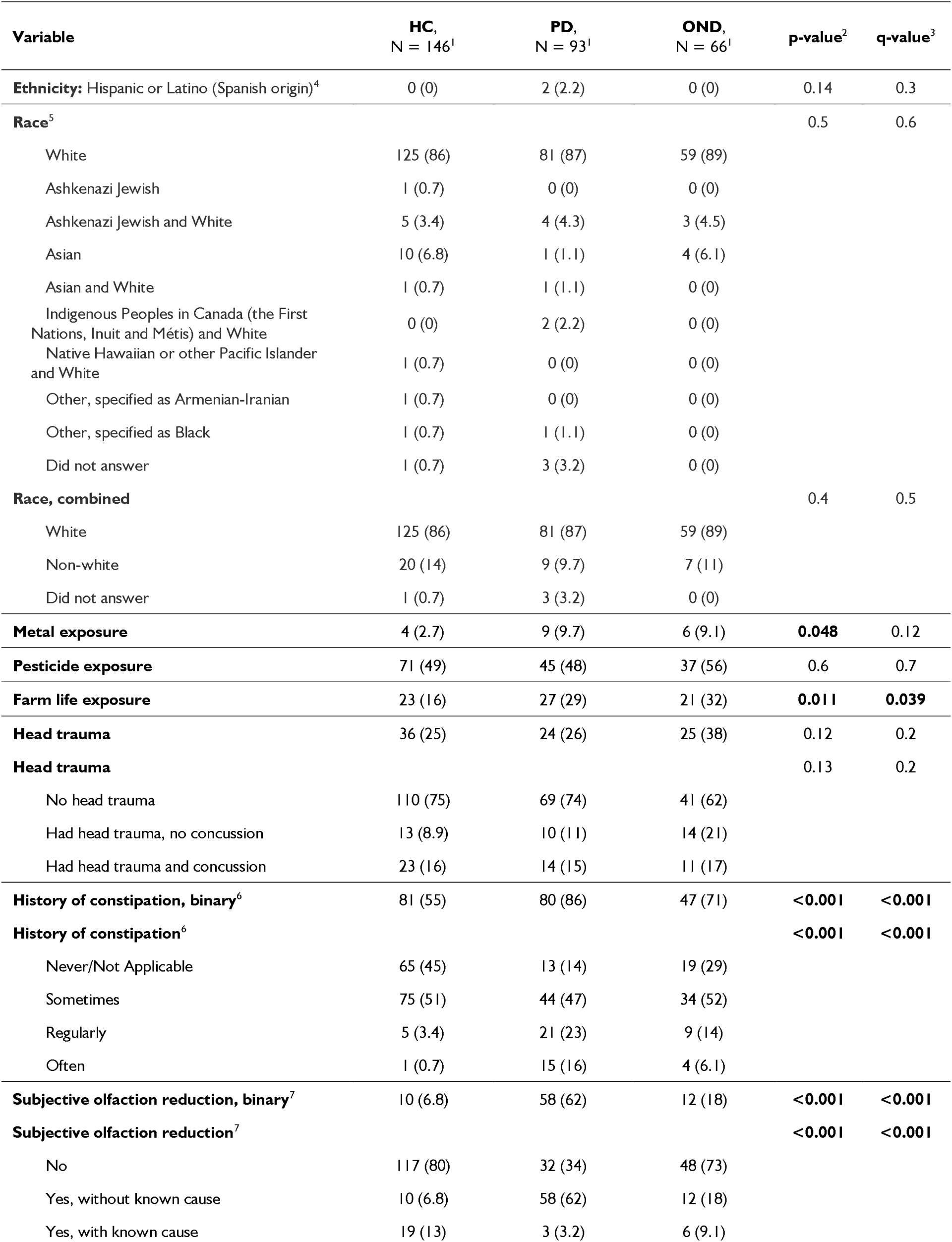
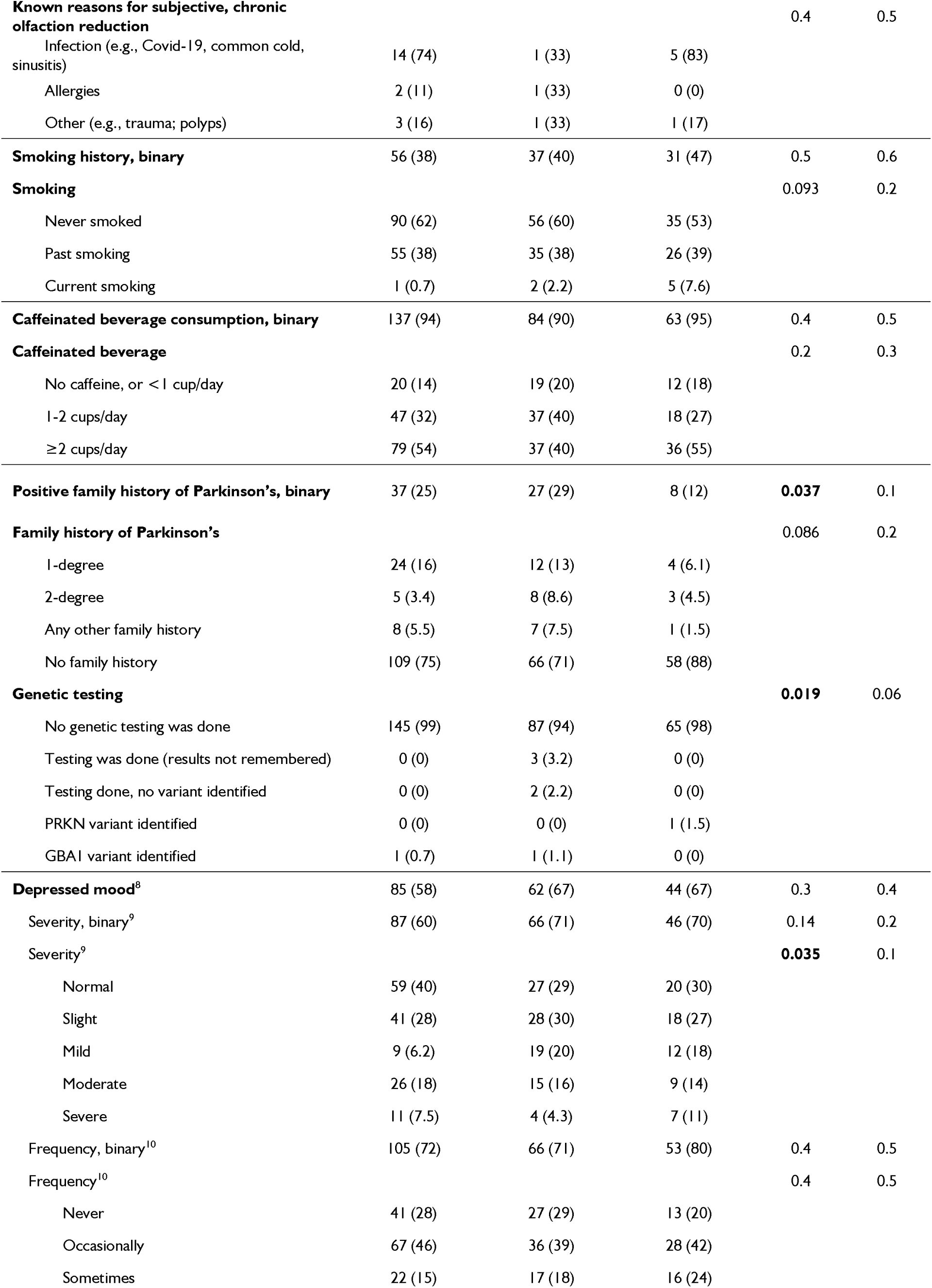
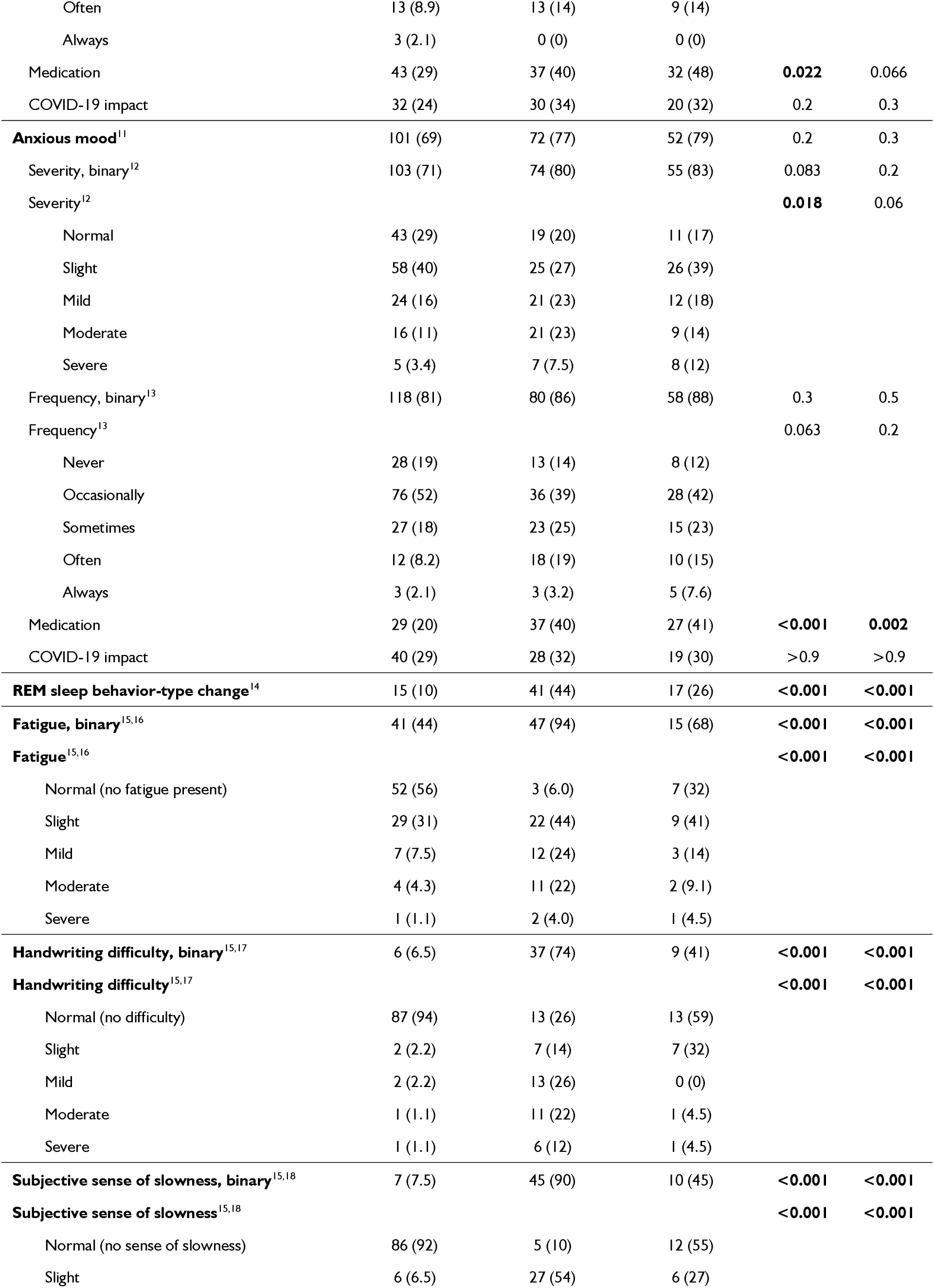
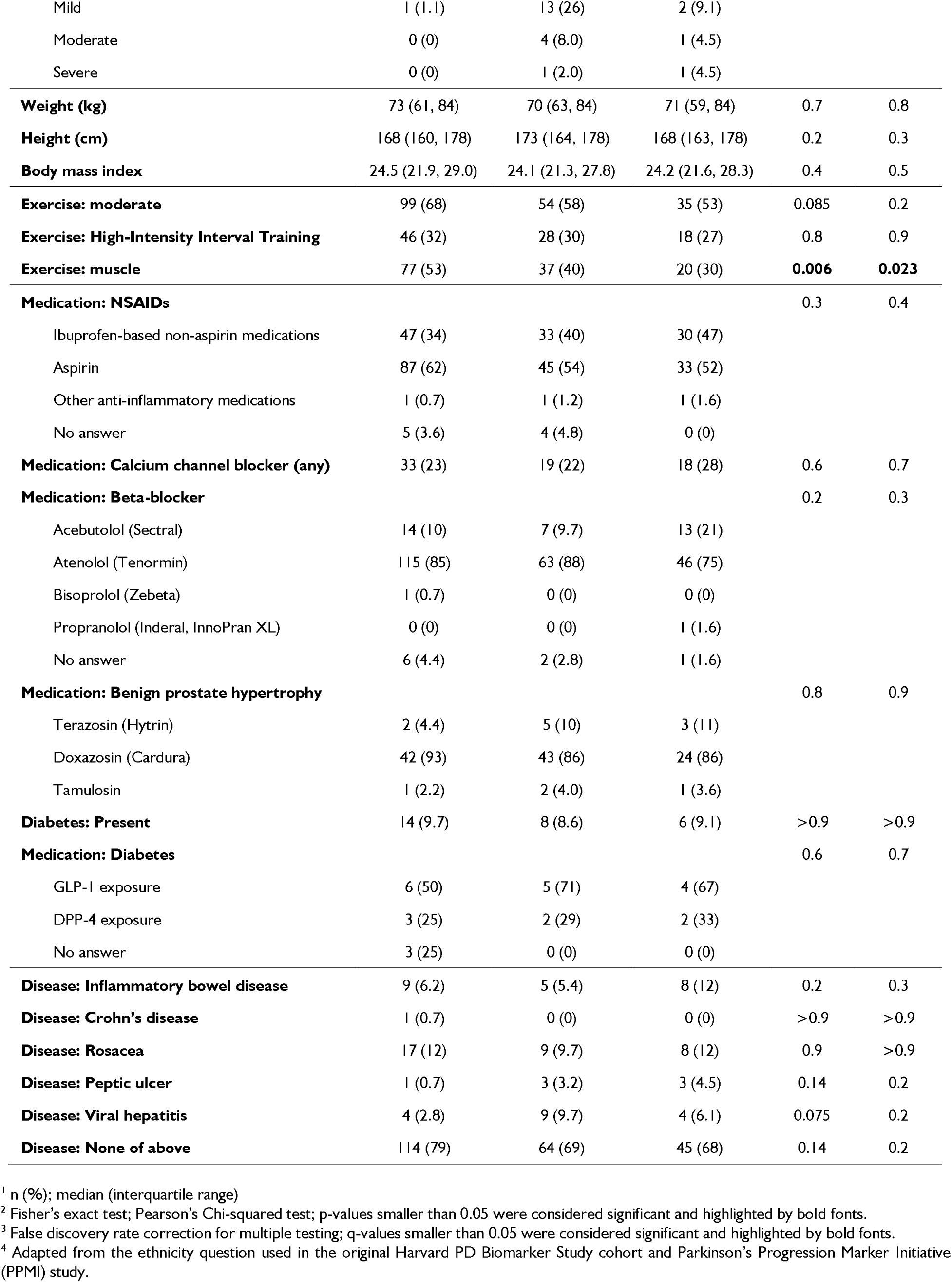
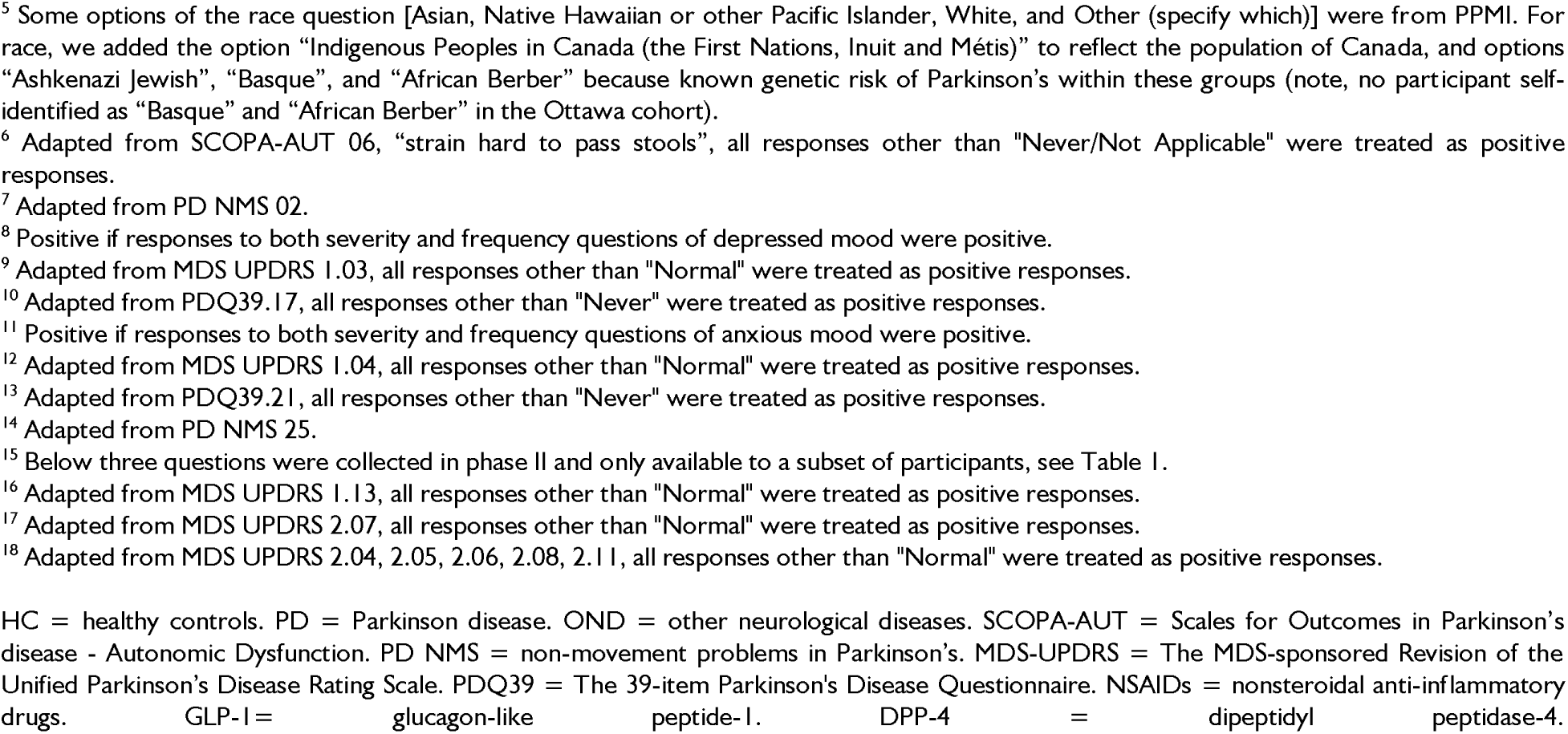
Complete list of variables collected in the Ottawa PD-RISK Index study using a self-administered, on-line questionnaire.

| Variable | HC,<br>N = 146<sup>1</sup> | PD,<br>N = 93<sup>1</sup> | OND,<br>N = 66<sup>1</sup> | p-value<sup>2</sup> | q-value<sup>3</sup> |
| --- | --- | --- | --- | --- | --- |
| **Ethnicity:** Hispanic or Latino (Spanish origin)<sup>4</sup> | 0 (0) | 2 (2.2) | 0 (0) | 0.14 | 0.3 |
| **Race<sup>5</sup>** |  |  |  | 0.5 | 0.6 |
| White | 125 (86) | 81 (87) | 59 (89) |  |  |
| Ashkenazi Jewish | 1 (0.7) | 0 (0) | 0 (0) |  |  |
| Ashkenazi Jewish and White | 5 (3.4) | 4 (4.3) | 3 (4.5) |  |  |
| Asian | 10 (6.8) | 1 (1.1) | 4 (6.1) |  |  |
| Asian and White | 1 (0.7) | 1 (1.1) | 0 (0) |  |  |
| Indigenous Peoples in Canada (the First Nations, Inuit and Métis) and White | 0 (0) | 2 (2.2) | 0 (0) |  |  |
| Native Hawaiian or other Pacific Islander and White | 1 (0.7) | 0 (0) | 0 (0) |  |  |
| Other, specified as Armenian-Iranian | 1 (0.7) | 0 (0) | 0 (0) |  |  |
| Other, specified as Black | 1 (0.7) | 1 (1.1) | 0 (0) |  |  |
| Did not answer | 1 (0.7) | 3 (3.2) | 0 (0) |  |  |
| **Race, combined** |  |  |  | 0.4 | 0.5 |
| White | 125 (86) | 81 (87) | 59 (89) |  |  |
| Non-white | 20 (14) | 9 (9.7) | 7 (11) |  |  |
| Did not answer | 1 (0.7) | 3 (3.2) | 0 (0) |  |  |
| **Metal exposure** | 4 (2.7) | 9 (9.7) | 6 (9.1) | **0.048** | 0.12 |
| **Pesticide exposure** | 71 (49) | 45 (48) | 37 (56) | 0.6 | 0.7 |
| **Farm life exposure** | 23 (16) | 27 (29) | 21 (32) | **0.011** | **0.039** |
| **Head trauma** | 36 (25) | 24 (26) | 25 (38) | 0.12 | 0.2 |
| **Head trauma** |  |  |  | 0.13 | 0.2 |
| No head trauma | 110 (75) | 69 (74) | 41 (62) |  |  |
| Had head trauma, no concussion | 13 (8.9) | 10 (11) | 14 (21) |  |  |
| Had head trauma and concussion | 23 (16) | 14 (15) | 11 (17) |  |  |
| **History of constipation, binary<sup>6</sup>** | 81 (55) | 80 (86) | 47 (71) | **<0.001** | **<0.001** |
| **History of constipation<sup>6</sup>** |  |  |  | **<0.001** | **<0.001** |
| Never/Not Applicable | 65 (45) | 13 (14) | 19 (29) |  |  |
| Sometimes | 75 (51) | 44 (47) | 34 (52) |  |  |
| Regularly | 5 (3.4) | 21 (23) | 9 (14) |  |  |
| Often | 1 (0.7) | 15 (16) | 4 (6.1) |  |  |
| **Subjective olfaction reduction, binary<sup>7</sup>** | 10 (6.8) | 58 (62) | 12 (18) | **<0.001** | **<0.001** |
| **Subjective olfaction reduction<sup>7</sup>** |  |  |  | **<0.001** | **<0.001** |
| No | 117 (80) | 32 (34) | 48 (73) |  |  |
| Yes, without known cause | 10 (6.8) | 58 (62) | 12 (18) |  |  |
| Yes, with known cause | 19 (13) | 3 (3.2) | 6 (9.1) |  |  |
| **Known reasons for subjective, chronic olfaction reduction** |  |  |  | 0.4 | 0.5 |
| Infection (e.g., Covid-19, common cold, sinusitis) | 14 (74) | 1 (33) | 5 (83) |  |  |
| Allergies | 2 (11) | 1 (33) | 0 (0) |  |  |
| Other (e.g., trauma; polyps) | 3 (16) | 1 (33) | 1 (17) |  |  |
| **Smoking history, binary** | 56 (38) | 37 (40) | 31 (47) | 0.5 | 0.6 |
| **Smoking** |  |  |  | 0.093 | 0.2 |
| Never smoked | 90 (62) | 56 (60) | 35 (53) |  |  |
| Past smoking | 55 (38) | 35 (38) | 26 (39) |  |  |
| Current smoking | 1 (0.7) | 2 (2.2) | 5 (7.6) |  |  |
| **Caffeinated beverage consumption, binary** | 137 (94) | 84 (90) | 63 (95) | 0.4 | 0.5 |
| **Caffeinated beverage** |  |  |  | 0.2 | 0.3 |
| No caffeine, or <1 cup/day | 20 (14) | 19 (20) | 12 (18) |  |  |
| 1-2 cups/day | 47 (32) | 37 (40) | 18 (27) |  |  |
| ≥2 cups/day | 79 (54) | 37 (40) | 36 (55) |  |  |
| **Positive family history of Parkinson's, binary** | 37 (25) | 27 (29) | 8 (12) | **0.037** | 0.1 |
| **Family history of Parkinson's** |  |  |  | 0.086 | 0.2 |
| 1-degree | 24 (16) | 12 (13) | 4 (6.1) |  |  |
| 2-degree | 5 (3.4) | 8 (8.6) | 3 (4.5) |  |  |
| Any other family history | 8 (5.5) | 7 (7.5) | 1 (1.5) |  |  |
| No family history | 109 (75) | 66 (71) | 58 (88) |  |  |
| **Genetic testing** |  |  |  | **0.019** | 0.06 |
| No genetic testing was done | 145 (99) | 87 (94) | 65 (98) |  |  |
| Testing was done (results not remembered) | 0 (0) | 3 (3.2) | 0 (0) |  |  |
| Testing done, no variant identified | 0 (0) | 2 (2.2) | 0 (0) |  |  |
| PRKN variant identified | 0 (0) | 0 (0) | 1 (1.5) |  |  |
| GBA1 variant identified | 1 (0.7) | 1 (1.1) | 0 (0) |  |  |
| **Depressed mood<sup>8</sup>** | 85 (58) | 62 (67) | 44 (67) | 0.3 | 0.4 |
| Severity, binary<sup>9</sup> | 87 (60) | 66 (71) | 46 (70) | 0.14 | 0.2 |
| Severity<sup>9</sup> |  |  |  | **0.035** | 0.1 |
| Normal | 59 (40) | 27 (29) | 20 (30) |  |  |
| Slight | 41 (28) | 28 (30) | 18 (27) |  |  |
| Mild | 9 (6.2) | 19 (20) | 12 (18) |  |  |
| Moderate | 26 (18) | 15 (16) | 9 (14) |  |  |
| Severe | 11 (7.5) | 4 (4.3) | 7 (11) |  |  |
| Frequency, binary<sup>10</sup> | 105 (72) | 66 (71) | 53 (80) | 0.4 | 0.5 |
| Frequency<sup>10</sup> |  |  |  | 0.4 | 0.5 |
| Never | 41 (28) | 27 (29) | 13 (20) |  |  |
| Occasionally | 67 (46) | 36 (39) | 28 (42) |  |  |
| Sometimes | 22 (15) | 17 (18) | 16 (24) |  |  |
| Often | 13 (8.9) | 13 (14) | 9 (14) |  |  |
| Always | 3 (2.1) | 0 (0) | 0 (0) |  |  |
| Medication | 43 (29) | 37 (40) | 32 (48) | **0.022** | 0.066 |
| COVID-19 impact | 32 (24) | 30 (34) | 20 (32) | 0.2 | 0.3 |
| **Anxious mood**<sup>11</sup> | 101 (69) | 72 (77) | 52 (79) | 0.2 | 0.3 |
| Severity, binary<sup>12</sup> | 103 (71) | 74 (80) | 55 (83) | 0.083 | 0.2 |
| Severity<sup>12</sup> |  |  |  | **0.018** | 0.06 |
| Normal | 43 (29) | 19 (20) | 11 (17) |  |  |
| Slight | 58 (40) | 25 (27) | 26 (39) |  |  |
| Mild | 24 (16) | 21 (23) | 12 (18) |  |  |
| Moderate | 16 (11) | 21 (23) | 9 (14) |  |  |
| Severe | 5 (3.4) | 7 (7.5) | 8 (12) |  |  |
| Frequency, binary<sup>13</sup> | 118 (81) | 80 (86) | 58 (88) | 0.3 | 0.5 |
| Frequency<sup>13</sup> |  |  |  | 0.063 | 0.2 |
| Never | 28 (19) | 13 (14) | 8 (12) |  |  |
| Occasionally | 76 (52) | 36 (39) | 28 (42) |  |  |
| Sometimes | 27 (18) | 23 (25) | 15 (23) |  |  |
| Often | 12 (8.2) | 18 (19) | 10 (15) |  |  |
| Always | 3 (2.1) | 3 (3.2) | 5 (7.6) |  |  |
| Medication | 29 (20) | 37 (40) | 27 (41) | **<0.001** | **0.002** |
| COVID-19 impact | 40 (29) | 28 (32) | 19 (30) | >0.9 | >0.9 |
| **REM sleep behavior-type change**<sup>14</sup> | 15 (10) | 41 (44) | 17 (26) | **<0.001** | **<0.001** |
| **Fatigue, binary**<sup>15,16</sup> | 41 (44) | 47 (94) | 15 (68) | **<0.001** | **<0.001** |
| **Fatigue**<sup>15,16</sup> |  |  |  | **<0.001** | **<0.001** |
| Normal (no fatigue present) | 52 (56) | 3 (6.0) | 7 (32) |  |  |
| Slight | 29 (31) | 22 (44) | 9 (41) |  |  |
| Mild | 7 (7.5) | 12 (24) | 3 (14) |  |  |
| Moderate | 4 (4.3) | 11 (22) | 2 (9.1) |  |  |
| Severe | 1 (1.1) | 2 (4.0) | 1 (4.5) |  |  |
| **Handwriting difficulty, binary**<sup>15,17</sup> | 6 (6.5) | 37 (74) | 9 (41) | **<0.001** | **<0.001** |
| **Handwriting difficulty**<sup>15,17</sup> |  |  |  | **<0.001** | **<0.001** |
| Normal (no difficulty) | 87 (94) | 13 (26) | 13 (59) |  |  |
| Slight | 2 (2.2) | 7 (14) | 7 (32) |  |  |
| Mild | 2 (2.2) | 13 (26) | 0 (0) |  |  |
| Moderate | 1 (1.1) | 11 (22) | 1 (4.5) |  |  |
| Severe | 1 (1.1) | 6 (12) | 1 (4.5) |  |  |
| **Subjective sense of slowness, binary**<sup>15,18</sup> | 7 (7.5) | 45 (90) | 10 (45) | **<0.001** | **<0.001** |
| **Subjective sense of slowness**<sup>15,18</sup> |  |  |  | **<0.001** | **<0.001** |
| Normal (no sense of slowness) | 86 (92) | 5 (10) | 12 (55) |  |  |
| Slight | 6 (6.5) | 27 (54) | 6 (27) |  |  |
| Mild | 1 (1.1) | 13 (26) | 2 (9.1) |  |  |
| Moderate | 0 (0) | 4 (8.0) | 1 (4.5) |  |  |
| Severe | 0 (0) | 1 (2.0) | 1 (4.5) |  |  |
| **Weight (kg)** | 73 (61, 84) | 70 (63, 84) | 71 (59, 84) | 0.7 | 0.8 |
| **Height (cm)** | 168 (160, 178) | 173 (164, 178) | 168 (163, 178) | 0.2 | 0.3 |
| **Body mass index** | 24.5 (21.9, 29.0) | 24.1 (21.3, 27.8) | 24.2 (21.6, 28.3) | 0.4 | 0.5 |
| **Exercise: moderate** | 99 (68) | 54 (58) | 35 (53) | 0.085 | 0.2 |
| **Exercise: High-Intensity Interval Training** | 46 (32) | 28 (30) | 18 (27) | 0.8 | 0.9 |
| **Exercise: muscle** | 77 (53) | 37 (40) | 20 (30) | **0.006** | **0.023** |
| **Medication: NSAIDs** |  |  |  | 0.3 | 0.4 |
| Ibuprofen-based non-aspirin medications | 47 (34) | 33 (40) | 30 (47) |  |  |
| Aspirin | 87 (62) | 45 (54) | 33 (52) |  |  |
| Other anti-inflammatory medications | 1 (0.7) | 1 (1.2) | 1 (1.6) |  |  |
| No answer | 5 (3.6) | 4 (4.8) | 0 (0) |  |  |
| **Medication: Calcium channel blocker (any)** | 33 (23) | 19 (22) | 18 (28) | 0.6 | 0.7 |
| **Medication: Beta-blocker** |  |  |  | 0.2 | 0.3 |
| Acebutolol (Sectral) | 14 (10) | 7 (9.7) | 13 (21) |  |  |
| Atenolol (Tenormin) | 115 (85) | 63 (88) | 46 (75) |  |  |
| Bisoprolol (Zebeta) | 1 (0.7) | 0 (0) | 0 (0) |  |  |
| Propranolol (Inderal, InnoPran XL) | 0 (0) | 0 (0) | 1 (1.6) |  |  |
| No answer | 6 (4.4) | 2 (2.8) | 1 (1.6) |  |  |
| **Medication: Benign prostate hypertrophy** |  |  |  | 0.8 | 0.9 |
| Terazosin (Hytrin) | 2 (4.4) | 5 (10) | 3 (11) |  |  |
| Doxazosin (Cardura) | 42 (93) | 43 (86) | 24 (86) |  |  |
| Tamulosin | 1 (2.2) | 2 (4.0) | 1 (3.6) |  |  |
| **Diabetes: Present** | 14 (9.7) | 8 (8.6) | 6 (9.1) | >0.9 | >0.9 |
| **Medication: Diabetes** |  |  |  | 0.6 | 0.7 |
| GLP-1 exposure | 6 (50) | 5 (71) | 4 (67) |  |  |
| DPP-4 exposure | 3 (25) | 2 (29) | 2 (33) |  |  |
| No answer | 3 (25) | 0 (0) | 0 (0) |  |  |
| **Disease: Inflammatory bowel disease** | 9 (6.2) | 5 (5.4) | 8 (12) | 0.2 | 0.3 |
| **Disease: Crohn's disease** | 1 (0.7) | 0 (0) | 0 (0) | >0.9 | >0.9 |
| **Disease: Rosacea** | 17 (12) | 9 (9.7) | 8 (12) | 0.9 | >0.9 |
| **Disease: Peptic ulcer** | 1 (0.7) | 3 (3.2) | 3 (4.5) | 0.14 | 0.2 |
| **Disease: Viral hepatitis** | 4 (2.8) | 9 (9.7) | 4 (6.1) | 0.075 | 0.2 |
| **Disease: None of above** | 114 (79) | 64 (69) | 45 (68) | 0.14 | 0.2 |

<sup>1</sup> n (%); median (interquartile range)
<sup>2</sup> Fisher's exact test; Pearson's Chi-squared test; p-values smaller than 0.05 were considered significant and highlighted by bold fonts.
<sup>3</sup> False discovery rate correction for multiple testing; q-values smaller than 0.05 were considered significant and highlighted by bold fonts.
<sup>4</sup> Adapted from the ethnicity question used in the original Harvard PD Biomarker Study cohort and Parkinson's Progression Marker Initiative (PPMI) study.
<sup>5</sup> Some options of the race question [Asian, Native Hawaiian or other Pacific Islander, White, and Other (specify which)] were from PPMI. For race, we added the option "Indigenous Peoples in Canada (the First Nations, Inuit and Métis)" to reflect the population of Canada, and options "Ashkenazi Jewish", "Basque", and "African Berber" because known genetic risk of Parkinson's within these groups (note, no participant self-identified as "Basque" and "African Berber" in the Ottawa cohort).
<sup>6</sup> Adapted from SCOPA-AUT 06, "strain hard to pass stools", all responses other than "Never/Not Applicable" were treated as positive responses.
<sup>7</sup> Adapted from PD NMS 02.
<sup>8</sup> Positive if responses to both severity and frequency questions of depressed mood were positive.
<sup>9</sup> Adapted from MDS UPDRS 1.03, all responses other than "Normal" were treated as positive responses.
<sup>10</sup> Adapted from PDQ39.17, all responses other than "Never" were treated as positive responses.
<sup>11</sup> Positive if responses to both severity and frequency questions of anxious mood were positive.
<sup>12</sup> Adapted from MDS UPDRS 1.04, all responses other than "Normal" were treated as positive responses.
<sup>13</sup> Adapted from PDQ39.21, all responses other than "Never" were treated as positive responses.
<sup>14</sup> Adapted from PD NMS 25.

**Supplementary Table 3:** Discriminative performances for each test in the entire PD-RISK Index study cohort.

| Test | Version | Median (interquartile range) |  |  | AUC (95% confidence interval) |  |  |
| --- | --- | --- | --- | --- | --- | --- | --- |
|  |  | HC | PD | OND | PD vs HC | PD vs OND | PD vs HC+OND |
| Scores, Questionnaire<sup>1</sup> | Original<sup>2</sup> | 50<br>(23–85) | 104<br>(71–148) | 77<br>(35–112) | 0.75<br>(0.69, 0.82) | 0.64<br>(0.56, 0.73) | 0.72<br>(0.66, 0.78) |
|  | Simplified<sup>3</sup> | 55<br>(29–82) | 210<br>(168–307) | 110<br>(44–198) | 0.96<br>(0.93, 0.98) | 0.75<br>(0.61, 0.89) | 0.92<br>(0.88, 0.96) |
| Scores, Smell Test | UPSIT<sup>4</sup> | 32<br>(29–35) | 17<br>(12–22) | 26<br>(18–33) | 0.93<br>(0.89, 0.96) | 0.73<br>(0.63, 0.83) | 0.87<br>(0.83, 0.92) |
|  | Simplified<sup>5</sup> | 7<br>(5–8) | 2<br>(1–4) | 4<br>(1–7) | 0.93<br>(0.89, 0.97) | 0.72<br>(0.57, 0.86) | 0.89<br>(0.84, 0.94) |
| **PD-RISK Index** | Original<sup>6</sup> | 7<br>(4–17) | 84<br>(60–95) | 28<br>(8–75) | 0.93<br>(0.9, 0.97) | 0.76<br>(0.67, 0.85) | 0.89<br>(0.84, 0.93) |
|  | **Simplified<sup>7</sup>** | **2**<br>**(1–7)** | **96**<br>**(69–100)** | **15**<br>**(2–91)** | **0.97**<br>**(0.95, 0.99)** | **0.77**<br>**(0.64, 0.9)** | **0.93**<br>**(0.89, 0.97)** |

<sup>1</sup> Formula to calculate the PD-RISK Index questionnaire score is listed in **Table 2**. For score ranges, the minimum score was based on a hypothetical 40-year-old female who responded negatively to all risk factors and positively to all protective factors (if applicable); the maximum score was based on a hypothetical 100-year-old male who responded positively to all risk factors and negatively to all protective factors (if applicable).
<sup>2</sup> Score range: 0.32–510 (see note 1). Actual score range in the PD-RISK Index study cohort: 0.384–323.7.
<sup>3</sup> Score range: 0.32–540 (see note 1). Actual score range in the PD-RISK Index study cohort: 0.384–398.4.
<sup>4</sup> Score range: 0–40, integer. Actual score range in the PD-RISK Index study cohort: 1–39.
<sup>5</sup> Score range: 0–8, integer. Actual score range in the PD-RISK Index study cohort: 0–8.
<sup>6</sup> Score range: 0–100. Actual score range in the PD-RISK Index study cohort: 0.94–99.89. Predicted probability x 100 based on logistic regression: $\text{logit}[p(\text{group} = \text{PD})] = 4.826 + 0.008 \times \text{original questionnaire score} - 0.243 \times \text{UPSIT score}.$
<sup>7</sup> Score range: 0–100. Actual score range within the PD-RISK Index study cohort: 0.17–99.99. Predicted probability x 100 based on logistic regression: $\text{logit}[p(\text{group} = \text{PD})] = -0.732 + 0.025 \times \text{simplified questionnaire score} - 0.709 \times \text{simplified smell test score}.$
This table compared score performances using the entire PD-RISK Index study cohort. HC = healthy controls. PD = Parkinson disease. OND = other neurological diagnoses. AUC = area under the receiver operating characteristic curve. UPSIT = University of Pennsylvania Smell Identification Test.

**Supplementary Table 4:**
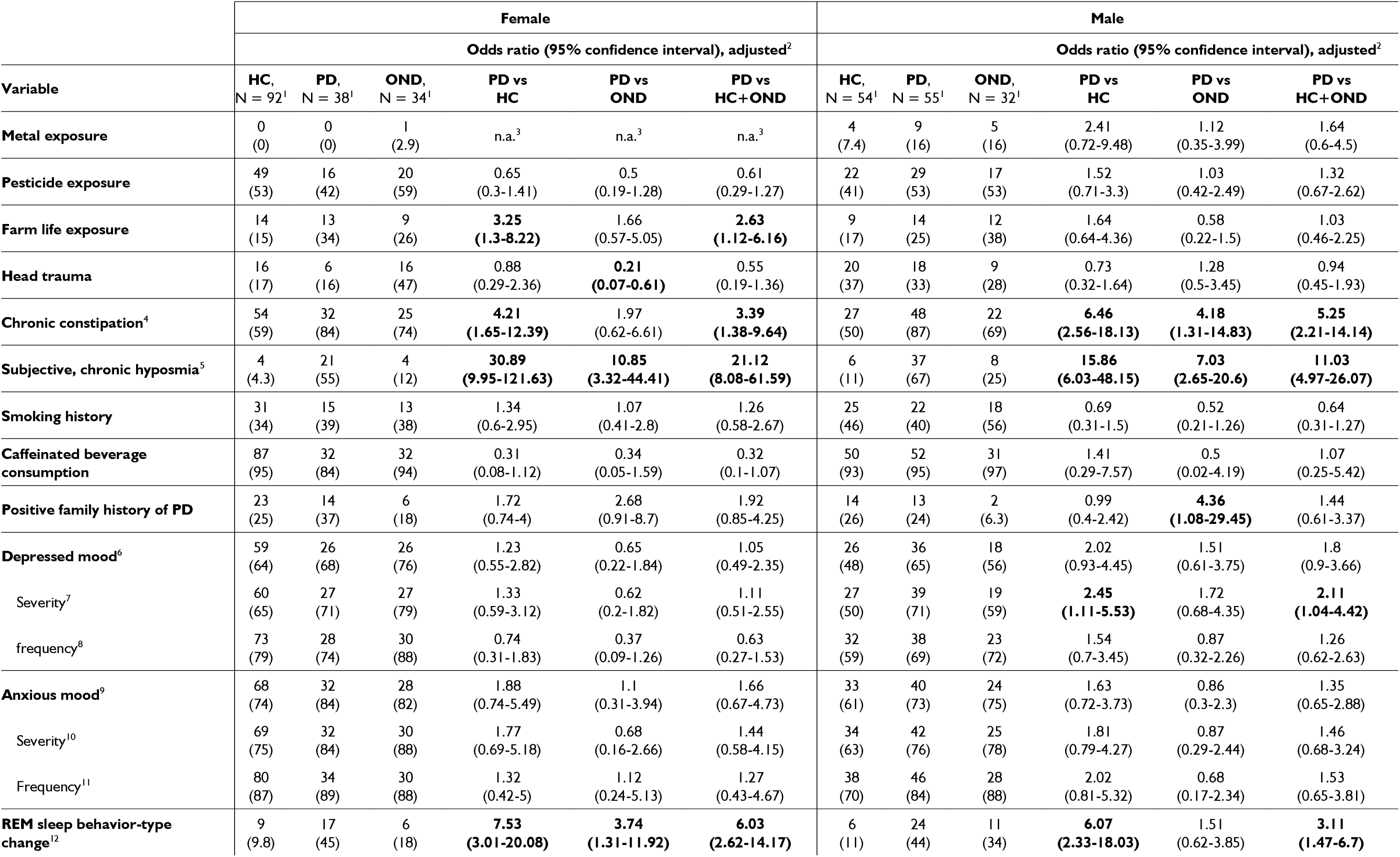
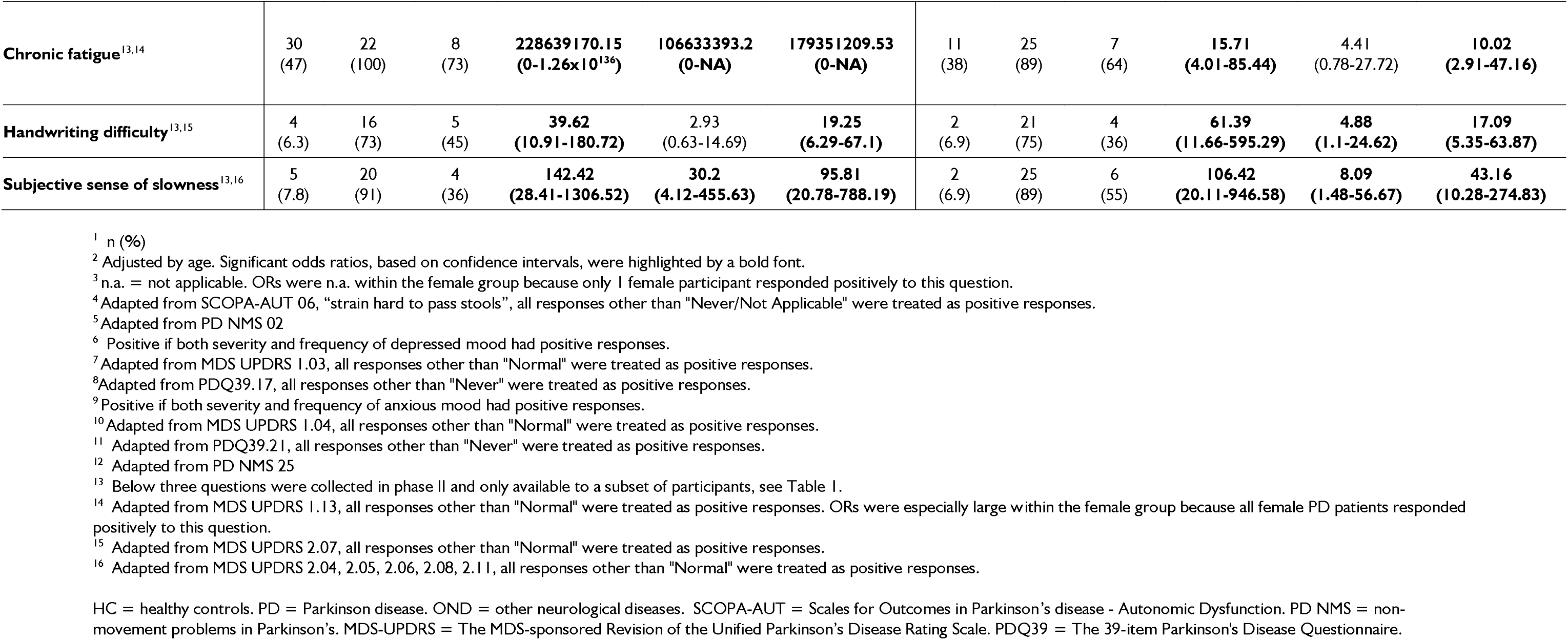
Summary of odds ratios for variables collected in the PD-RISK Index questionnaire, *listed by sex*.

**Supplementary Table 5:**
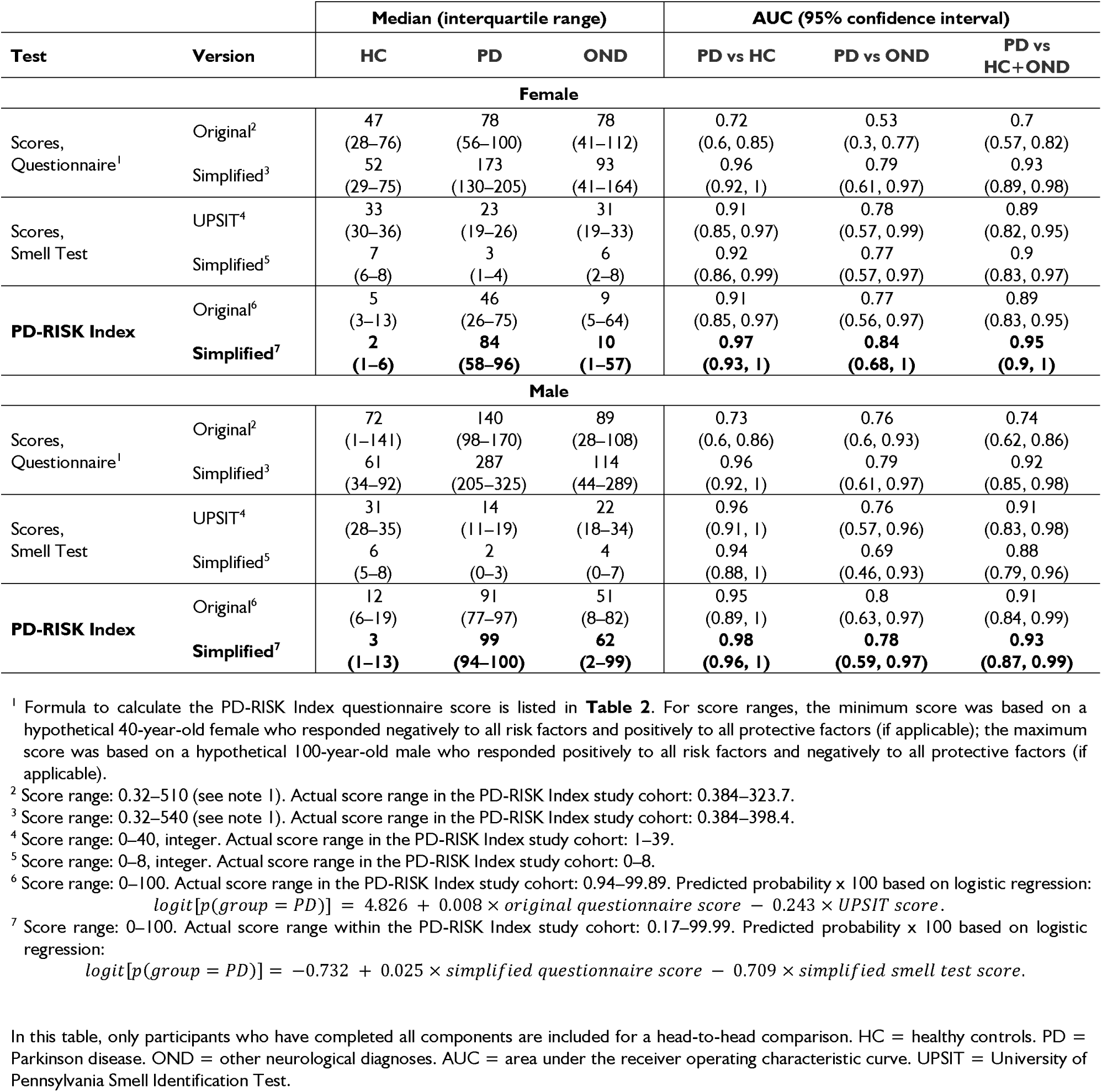
Discriminative performances for each test score in participants that completed all PD-RISK Index study assessments, *listed by sex*.

|  |  | Median (interquartile range) |  |  | AUC (95% confidence interval) |  |  |
| --- | --- | --- | --- | --- | --- | --- | --- |
| Test | Version | HC | PD | OND | PD vs HC | PD vs OND | PD vs HC+OND |
| Female |  |  |  |  |  |  |  |
| Scores, Questionnaire<sup>1</sup> | Original<sup>2</sup> | 47<br>(28–76) | 78<br>(56–100) | 78<br>(41–112) | 0.72<br>(0.6, 0.85) | 0.53<br>(0.3, 0.77) | 0.7<br>(0.57, 0.82) |
|  | Simplified<sup>3</sup> | 52<br>(29–75) | 173<br>(130–205) | 93<br>(41–164) | 0.96<br>(0.92, 1) | 0.79<br>(0.61, 0.97) | 0.93<br>(0.89, 0.98) |
| Scores, Smell Test | UPSIT<sup>4</sup> | 33<br>(30–36) | 23<br>(19–26) | 31<br>(19–33) | 0.91<br>(0.85, 0.97) | 0.78<br>(0.57, 0.99) | 0.89<br>(0.82, 0.95) |
|  | Simplified<sup>5</sup> | 7<br>(6–8) | 3<br>(1–4) | 6<br>(2–8) | 0.92<br>(0.86, 0.99) | 0.77<br>(0.57, 0.97) | 0.9<br>(0.83, 0.97) |
| PD-RISK Index | Original<sup>6</sup> | 5<br>(3–13) | 46<br>(26–75) | 9<br>(5–64) | 0.91<br>(0.85, 0.97) | 0.77<br>(0.56, 0.97) | 0.89<br>(0.83, 0.95) |
|  | Simplified<sup>7</sup> | 2<br>(1–6) | 84<br>(58–96) | 10<br>(1–57) | 0.97<br>(0.93, 1) | 0.84<br>(0.68, 1) | 0.95<br>(0.9, 1) |
| Male |  |  |  |  |  |  |  |
| Scores, Questionnaire<sup>1</sup> | Original<sup>2</sup> | 72<br>(1–141) | 140<br>(98–170) | 89<br>(28–108) | 0.73<br>(0.6, 0.86) | 0.76<br>(0.6, 0.93) | 0.74<br>(0.62, 0.86) |
|  | Simplified<sup>3</sup> | 61<br>(34–92) | 287<br>(205–325) | 114<br>(44–289) | 0.96<br>(0.92, 1) | 0.79<br>(0.61, 0.97) | 0.92<br>(0.85, 0.98) |
| Scores, Smell Test | UPSIT<sup>4</sup> | 31<br>(28–35) | 14<br>(11–19) | 22<br>(18–34) | 0.96<br>(0.91, 1) | 0.76<br>(0.57, 0.96) | 0.91<br>(0.83, 0.98) |
|  | Simplified<sup>5</sup> | 6<br>(5–8) | 2<br>(0–3) | 4<br>(0–7) | 0.94<br>(0.88, 1) | 0.69<br>(0.46, 0.93) | 0.88<br>(0.79, 0.96) |
| PD-RISK Index | Original<sup>6</sup> | 12<br>(6–19) | 91<br>(77–97) | 51<br>(8–82) | 0.95<br>(0.89, 1) | 0.8<br>(0.63, 0.97) | 0.91<br>(0.84, 0.99) |
|  | Simplified<sup>7</sup> | 3<br>(1–13) | 99<br>(94–100) | 62<br>(2–99) | 0.98<br>(0.96, 1) | 0.78<br>(0.59, 0.97) | 0.93<br>(0.87, 0.99) |

<sup>1</sup> Formula to calculate the PD-RISK Index questionnaire score is listed in **Table 2**. For score ranges, the minimum score was based on a hypothetical 40-year-old female who responded negatively to all risk factors and positively to all protective factors (if applicable); the maximum score was based on a hypothetical 100-year-old male who responded positively to all risk factors and negatively to all protective factors (if applicable).
<sup>2</sup> Score range: 0.32–510 (see note 1). Actual score range in the PD-RISK Index study cohort: 0.384–323.7.
<sup>3</sup> Score range: 0.32–540 (see note 1). Actual score range in the PD-RISK Index study cohort: 0.384–398.4.
<sup>4</sup> Score range: 0–40, integer. Actual score range in the PD-RISK Index study cohort: 1–39.
<sup>5</sup> Score range: 0–8, integer. Actual score range in the PD-RISK Index study cohort: 0–8.
<sup>6</sup> Score range: 0–100. Actual score range in the PD-RISK Index study cohort: 0.94–99.89. Predicted probability x 100 based on logistic regression: $\text{logit}[p(\text{group} = \text{PD})] = 4.826 + 0.008 \times \text{original questionnaire score} - 0.243 \times \text{UPSIT score}.$
<sup>7</sup> Score range: 0–100. Actual score range within the PD-RISK Index study cohort: 0.17–99.99. Predicted probability x 100 based on logistic regression: $\text{logit}[p(\text{group} = \text{PD})] = -0.732 + 0.025 \times \text{simplified questionnaire score} - 0.709 \times \text{simplified smell test score}.$
In this table, only participants who have completed all components are included for a head-to-head comparison. HC = healthy controls. PD = Parkinson disease. OND = other neurological diagnoses. AUC = area under the receiver operating characteristic curve. UPSIT = University of Pennsylvania Smell Identification Test.

**Supplementary Table 6:** Performance metrics for each test score and thresholds regarding diagnostic classification, listed for *female* participants.

| Test | Version | Overall threshold |  |  |  |  |  |  | Sex-specific threshold |  |  |  |  |  |  |
| --- | --- | --- | --- | --- | --- | --- | --- | --- | --- | --- | --- | --- | --- | --- | --- |
|  |  | Threshold<sup>1</sup> | Sensitivity | Specificity | LR+ | LR- | PPV | NPV | Threshold<sup>1</sup> | Sensitivity | Specificity | LR+ | LR- | PPV | NPV |
| PD vs HC+OND |  |  |  |  |  |  |  |  |  |  |  |  |  |  |  |
| Scores, Questionnaire | Original | ≥ 95.83 | 0.38 | 0.91 | 4.08 | 0.68 | 0.53 | 0.84 | ≥ 55 | 0.81 | 0.56 | 1.84 | 0.34 | 0.34 | 0.91 |
|  | Simplified | ≥ 93.3 | 0.95 | 0.77 | 4.2 | 0.06 | 0.54 | 0.98 | ≥ 90.7 | 1 | 0.76 | 4.17 | 0 | 0.54 | 1 |
| Scores, Smell Test | UPSIT | ≤ 28.5 | 1 | 0.77 | 4.41 | 0 | 0.55 | 1 | ≤ 28.5 | 1 | 0.77 | 4.41 | 0 | 0.55 | 1 |
|  | Simplified | ≤ 4.5 | 0.86 | 0.83 | 4.95 | 0.17 | 0.58 | 0.95 | ≤ 5.5 | 0.95 | 0.73 | 3.57 | 0.06 | 0.5 | 0.98 |
| PD-RISK Index | Original | ≥ 15.67 | 1 | 0.76 | 4.17 | 0 | 0.54 | 1 | ≥ 15.67 | 1 | 0.76 | 4.17 | 0 | 0.54 | 1 |
|  | Simplified | ≥ 22.72 | 0.95 | 0.89 | 8.93 | 0.05 | 0.71 | 0.99 | ≥ 33.1 | 0.95 | 0.91 | 10.2 | 0.05 | 0.74 | 0.99 |
| PD vs HC |  |  |  |  |  |  |  |  |  |  |  |  |  |  |  |
| Scores, Questionnaire | Original | ≥ 95.83 | 0.38 | 0.94 | 6.1 | 0.66 | 0.53 | 0.74 | ≥ 55 | 0.81 | 0.58 | 1.92 | 0.33 | 0.34 | 0.8 |
|  | Simplified | ≥ 90.9 | 1 | 0.81 | 5.33 | 0 | 0.54 | 0.91 | ≥ 90.7 | 1 | 0.81 | 5.33 | 0 | 0.54 | 0.91 |
| Scores, Smell Test | UPSIT | ≤ 28.5 | 1 | 0.8 | 4.92 | 0 | 0.55 | 0.88 | ≤ 28.5 | 1 | 0.8 | 4.92 | 0 | 0.55 | 0.88 |
|  | Simplified | ≤ 4.5 | 0.86 | 0.86 | 6.1 | 0.17 | 0.58 | 0.85 | ≤ 4.5 | 0.86 | 0.86 | 6.1 | 0.17 | 0.58 | 0.85 |
| PD-RISK Index | Original | ≥ 15.67 | 1 | 0.8 | 4.92 | 0 | 0.54 | 0.89 | ≥ 15.67 | 1 | 0.8 | 4.92 | 0 | 0.54 | 0.89 |
|  | Simplified | ≥ 22.72 | 0.95 | 0.92 | 12.19 | 0.05 | 0.71 | 0.87 | ≥ 33.1 | 0.95 | 0.94 | 15.24 | 0.05 | 0.74 | 0.87 |
| PD vs OND |  |  |  |  |  |  |  |  |  |  |  |  |  |  |  |
| Scores, Questionnaire | Original | ≥ 90.1 | 0.43 | 0.73 | 1.57 | 0.79 | 0.47 | 0.1 | ≥ 52 | 0.81 | 0.45 | 1.48 | 0.42 | 0.33 | 0.11 |
|  | Simplified | ≥ 93.3 | 0.95 | 0.55 | 2.1 | 0.09 | 0.54 | 0.1 | ≥ 94.4 | 0.95 | 0.55 | 2.1 | 0.09 | 0.56 | 0.1 |
| Scores, Smell Test | UPSIT | ≤ 28.5 | 1 | 0.64 | 2.75 | 0 | 0.55 | 0.12 | ≤ 28.5 | 1 | 0.64 | 2.75 | 0 | 0.55 | 0.12 |
|  | Simplified | ≤ 5.5 | 0.95 | 0.64 | 2.62 | 0.07 | 0.5 | 0.12 | ≤ 5.5 | 0.95 | 0.64 | 2.62 | 0.07 | 0.5 | 0.12 |
| PD-RISK Index | Original | ≥ 25.62 | 0.76 | 0.73 | 2.79 | 0.33 | 0.57 | 0.12 | ≥ 12.72 | 1 | 0.55 | 2.2 | 0 | 0.5 | 0.11 |
|  | Simplified | ≥ 19.56 | 0.95 | 0.73 | 3.49 | 0.07 | 0.69 | 0.12 | ≥ 26.87 | 0.95 | 0.73 | 3.49 | 0.07 | 0.71 | 0.12 |

<sup>1</sup> Optimal thresholds are based on the maximum Youden Index value. Signs (≥ or ≤) indicate classification of having Parkinson's.

**Supplementary Table 7:**
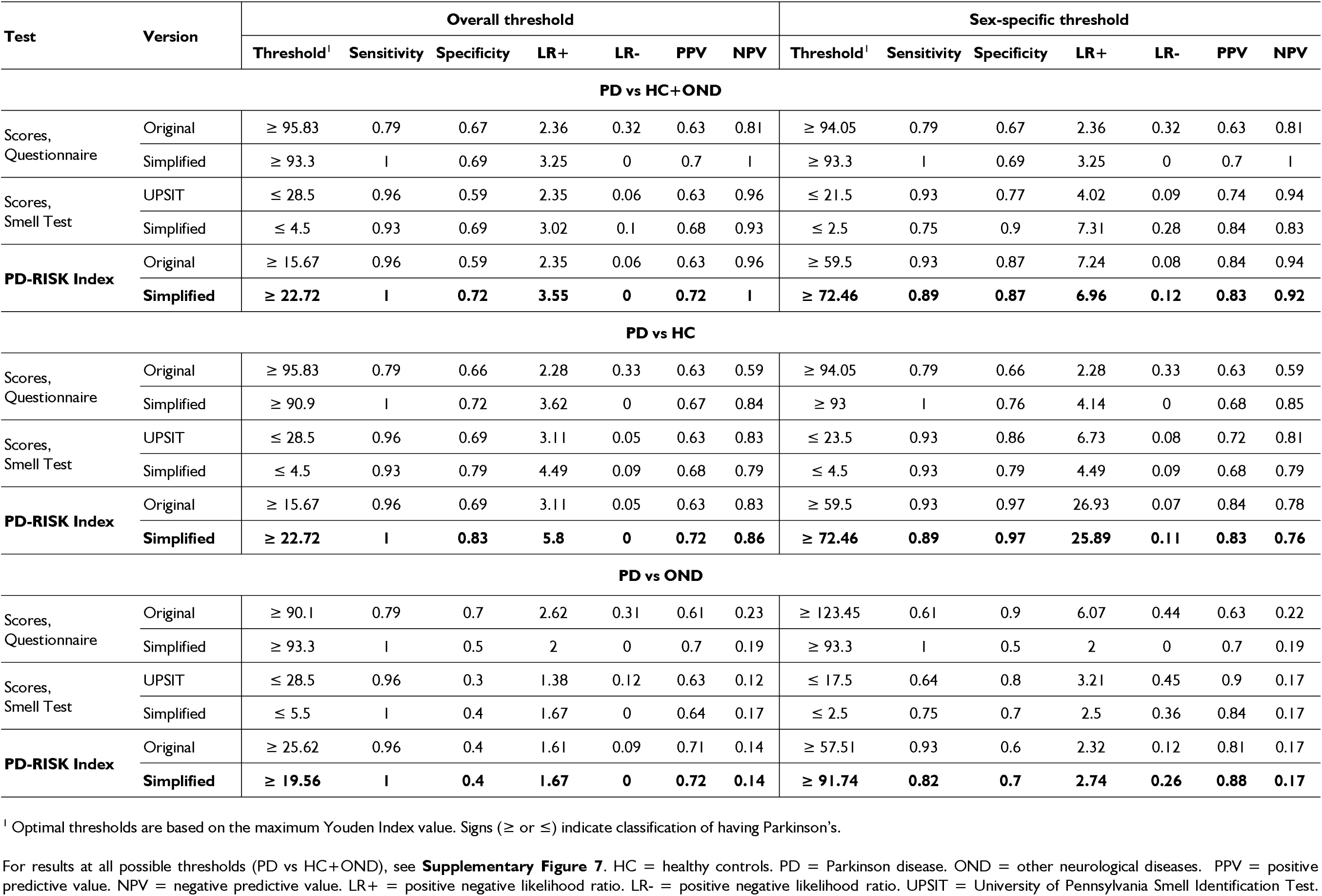
Performance metrics for each test score and thresholds regarding diagnostic classification, listed for *male* participants.

**Supplementary Table 8:**
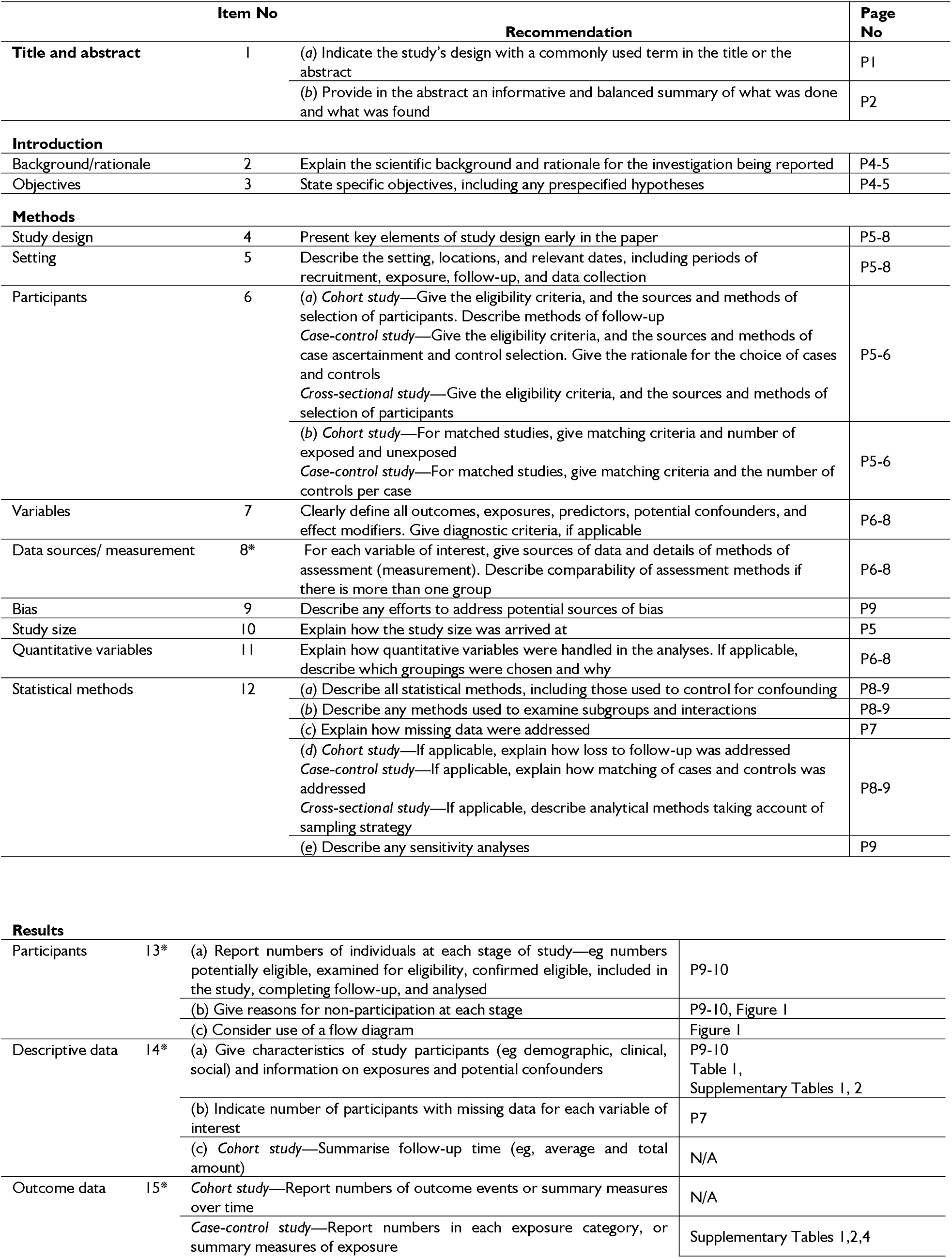
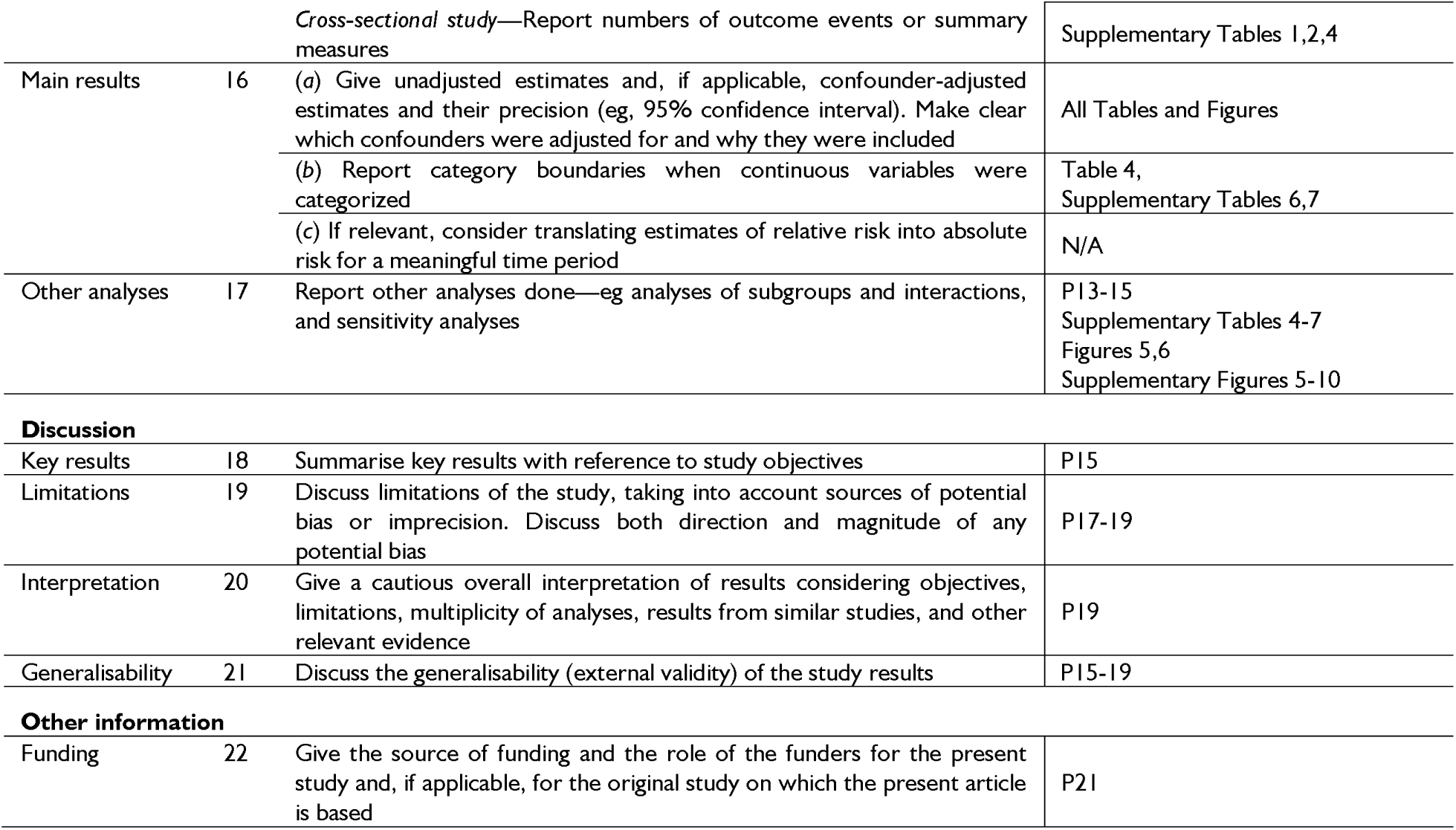
STROBE Statement - checklist of items in a report of observational studies.

**Supplementary Table 9:** Standards for Reporting of Diagnostic Accuracy Studies (STARD) checklist.

| Section & Topic | No | Item | Reported on page # |
| --- | --- | --- | --- |
| **TITLE OR ABSTRACT** |  |  |  |
|  | 1 | Identification as a study of diagnostic accuracy using at least one measure of accuracy (such as sensitivity, specificity, predictive values, or AUC) | P1 |
| **ABSTRACT** |  |  |  |
|  | 2 | Structured summary of study design, methods, results, and conclusions (for specific guidance, STARD for Abstracts) | P2 |
| **INTRODUCTION** |  |  |  |
|  | 3 | Scientific and clinical background, including the intended use and clinical role of the index test | P4-5 |
|  | 4 | Study objectives and hypotheses | P4-5 |
| **METHODS** |  |  |  |
| *Study design* | 5 | Whether data collection was planned before the index test and reference standard were performed (prospective study) or after (retrospective study) | P5-6 |
| *Participants* | 6 | Eligibility criteria | P5-6 |
|  | 7 | On what basis potentially eligible participants were identified (such as symptoms, results from previous tests, inclusion in registry) | P5-6 |
|  | 8 | Where and when potentially eligible participants were identified (setting, location and dates) | P5-6 |
|  | 9 | Whether participants formed a consecutive, random or convenience series | P5-6 |
| *Test methods* | 10a | Index test, in sufficient detail to allow replication | P6-7, Table 2 |
|  | 10b | Reference standard, in sufficient detail to allow replication | P6 |
|  | 11 | Rationale for choosing the reference standard (if alternatives exist) | P6 |
|  | 12a | Definition of and rationale for test positivity cut-offs or result categories of the index test, distinguishing pre-specified from exploratory | P8-9 |
|  | 12b | Definition of and rationale for test positivity cut-offs or result categories of the reference standard, distinguishing pre-specified from exploratory | P6 |
|  | 13a | Whether clinical information and reference standard results were available to the performers/readers of the index test | N/A. Same scoring scheme automatically applied to all participants. |
|  | 13b | Whether clinical information and index test results were available to the assessors of the reference standard | P6 |
| *Analysis* | 14 | Methods for estimating or comparing measures of diagnostic accuracy | P8-9 |
|  | 15 | How indeterminate index test or reference standard results were handled | P8-9 |
|  | 16 | How missing data on the index test and reference standard were handled | P7 |
|  | 17 | Any analyses of variability in diagnostic accuracy, distinguishing pre-specified from exploratory | P8-9 |
|  | 18 | Intended sample size and how it was determined | P5 |
| **RESULTS** |  |  |  |
| *Participants* | 19 | Flow of participants, using a diagram | Figure 1 |
|  | 20 | Baseline demographic and clinical characteristics of participants | P9-10, Table 1 |
|  | 21a | Distribution of severity of disease in those with the target condition | P5; Table 1 |
|  | 21b | Distribution of alternative diagnoses in those without the target condition | P5, P9; Table 1 |
|  | 22 | Time interval and any clinical interventions between index test and reference standard | P6 |
| *Test results* | 23 | Cross tabulation of the index test results (or their distribution) by the results of the reference standard | Figures 2, 3 |
|  | 24 | Estimates of diagnostic accuracy and their precision (such as 95% confidence intervals) | Figures 2, 3; Supplementary Figures 2-8; Tables 3, 4; Supplementary Tables 3, 5-7 |
|  | 25 | Any adverse events from performing the index test or the reference standard | P10 |
| **DISCUSSION** |  |  |  |
|  | **26** | Study limitations, including sources of potential bias, statistical uncertainty, and generalisability | P17-19 |
|  | **27** | Implications for practice, including the intended use and clinical role of the index test | P19 |
| **OTHER INFORMATION** |  |  |  |
|  | **28** | Registration number and name of registry | N/A; observational study |
|  | **29** | Where the full study protocol can be accessed | P22 |
|  | **30** | Sources of funding and other support; role of funders | P21 |

